# Cholangiocyte glycocalyx degradation boosts primary sclerosing cholangitis

**DOI:** 10.1101/2024.06.27.24309484

**Authors:** Friederike Klein, Freya Wellhöner, Anika Freise, Kristina M. Niculovic, Howard Junca, Manuel Vicente, Elina Kats, dos Anjos Borges Luiz Gustavo, Leonard Knegendorf, Karsten Cirksena, Antonia Marie Triefenbach, Franziska Woelfl, Helin Fatma Abdullah, Meike Schulz, Iris Plumeier, Silke Kahl, Iris Albers, Martijn Zoodsma, Marius Vital, Torsten Voigtländer, Henrike Lenzen, Jessica Schmitz, Anna Saborowski, Michael P. Manns, Philipp Solbach, Jan Hinrich Bräsen, Gisa Gerold, Cheng-Jian Xu, Heiner Wedemeyer, Anja K. Münster-Kühnel, Dietmar H. Pieper, Benjamin Heidrich

**Author notes:** These authors contributed equally to this work (Friederike Klein, Freya Wellhöner, Anika Freise).

## Abstract

**Primary sclerosing cholangitis (PSC) is an inflammatory disease of the biliary tract eventually leading to bile duct destruction, liver failure, cholangiocellular adenocarcinoma and/or death. No disease modifying treatments are available**^1^**. Especially cytotoxicity of bile acids, are discussed as potential driver of disease progression**^2^**. Cholangiocytes are protected by a bicarbonate umbrella formed by the glycocalyx, a dense layer of membrane bound polyglycans extending into the extracellular space**^3,4^**. Bile of PSC patients harbors a unique microbiome**^5^**. Here we identified a new factor in the pathogenesis of PSC. The bacterial degradation of sialic acid and galactose are associated with a poor event free survival of PSC patients and could identify bacterial liberation of sialic acid as crucial element in cholangiocyte damage using cell culture experiments, individualized organoid models and liver biopsies. With this study the view on bacteria-host interactions in bile duct associated diseases is widened. Functional patterns of the bacterial community are crucial for bile duct destruction in PSC patients. This opens a new field of diagnostic tools, disease modifying treatment options and identification of patients at risk.**

## Introduction

Primary sclerosing cholangitis (PSC) is an inflammatory disease of the biliary tract. The clinical manifestation is characterized by strictures of the bile duct leading to cholestasis and recurring bacterial cholangitis^6^. The clinical course of disease is highly variable, varying from mild to severe with the development of end-stage liver disease, cholangiocellular adenocarcinoma (CCA) and/or death in early adult life^7^. No disease modifying treatment is available so far. Therapeutic options limited to the treatment of complications like administration of antibiotics in terms of bacterial cholangitis or endoscopic interventions via endoscopic retrograde cholangiography (ERC) in terms of biliary strictures and cholestasis^1^.

The pathogenesis is widely unknown and thought to be multifactorial^2^. Bile acid toxicity against cholangiocytes, the epithelial cells lining the luminal side of bile ducts, is one of these accepted factors contributing to inflammation and destruction of the biliary tract^4,8^. Bile was long thought to be sterile. Meanwhile, the presence of a biliary microbiome is well accepted and the structure of the bacterial community in bile of patients suffering from PSC shows distinct patterns compared to patients without a PSC^5^. Former studies showed an improvement of biochemical disease activity after the administration of antibiotics in a subset of patients. Furthermore, one study evaluated the use of fecal microbiome transfer in patients with PSC. In line with the studies on the use of antibiotics, FMT led to an improvement in a subgroup of patients^9^. These, studies indicate an important role of bacteria in the pathogenesis of PSC. However, it needs to be clarified which patients might have a benefit of therapies targeting the biliary and/or intestinal microbiota. Therefore, we aim to understand the role of bacteria of the biliary microbiome in the pathogenesis of PSC, to identify patterns within the bacterial community that correlate with severe courses of disease and to find new target options for an individualized therapy.

## Results

### Identification of bacterial communities in bile

The course of disease observed in patients with primary sclerosing cholangitis (PSC) is known to be highly variable varying from mild expressions to severe complications in early live^7^. In order to find corresponding variable signatures in the biliary microbial community of affected patients, the 16S rRNA gene profiles of 209 bile samples from 117 different patients suffering from PSC were analyzed. *Enterococcus* and *Streptococcus* were observed in all samples at a high mean relative abundance >10 % each (Extended DataTable 1). Further prominent genera with a mean relative abundance of >1 % and a prevalence >80 % were anaerobic bacteria such as *Schaalia, Bifidobacterium, Prevotella* and *Bacteroides* as well as bacteria of the Enterobacteriaceae family and *Staphylococcus, Haemophilus* and *Granulicatella* (Extended Data Table 1). *Clostridium sensu stricto* and *Pseudomonas* showed a similarly high mean relative abundance of >1 % but a prevalence <80 % (Extended Data Table 1). Importantly, the 16S profiles in the majority of bile samples (n = 171, 82 %) were dominated by a single bacterial genus (dominance = relative abundance >33 %) (Figure 1A). Similar to other body sites with a low bacterial load microbiome, multiple genera can be the dominating one in different bile samples. Three genera, i.e. *Enterococcus* (n = 60), *Streptococcus* (n = 29) and *Staphylococcus* (n = 12) were often dominating in bile and respective samples were termed bilotype (BT). As genera of the family Enterobacteriaceae, which were also dominating in a high amount of samples (n = 38), cannot be differentiated at high accuracy based on 16S rRNA gene sequences only^10^, these samples were termed as Enterobacteriaceae BT. Twenty-two samples were dominated by other less prevalent genera whereas 38 samples were not dominated by any bacterial genus (no BT). The presence of different BTs in bile were similarly observed in an independent PSC cohort (Extended Data Figure 1).

**Figure 1:**
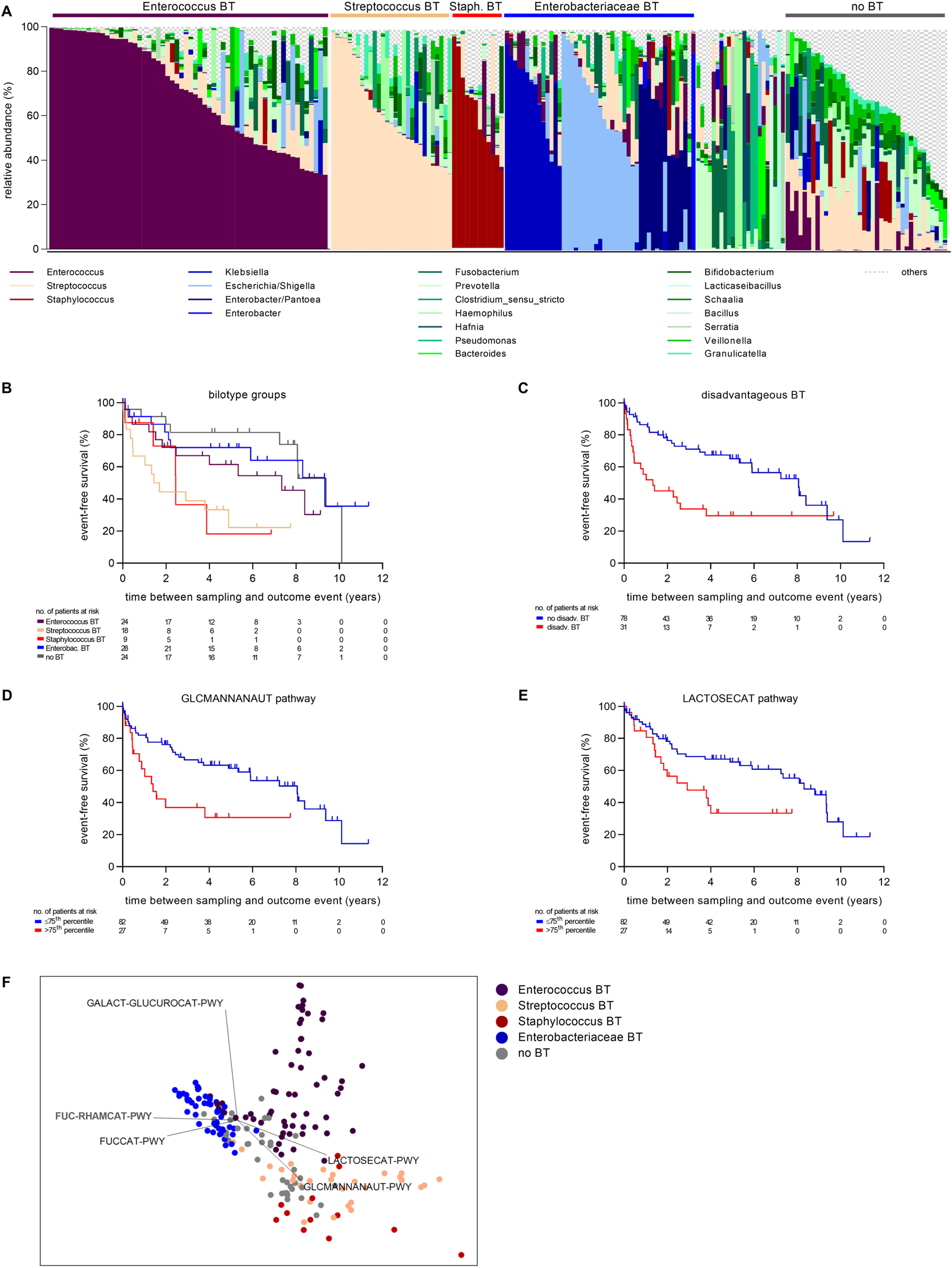
Correlation of clinical course of PSC with patterns in the biliary microbiome. **A** 16S rRNA gene profile of all bile samples. Each bar indicates one sample. Genera with a mean relative abundance of less than 1% are subsumed as others. n = 209. **B-E** Event-free survival of PSC patients. Outcome defining event: liver transplantation, cholangiocellular adenocarcinoma (CCA) and death. Excluded were all patients with an existing CCA at the time point of sampling (included n = 109 patients) **B** According to the bilotype. Streptococcus BT vs. no BT p = 5.9-4. Staphylococcus BT vs no BT p = 0.008. Streptococcus vs Enterobacteriaceae BT p=0.006. Enterobact. = Enterobacteriaceae, BT = bilotype. **C** According to disadvantageous BT in patient history. p = 0.001. disadv. = disadvantageous, BT = bilotype. **D and E** According to relative abundance of GLCMANNANAUT pathway (p = 0.006) or LACTOSECAT pathway (p = 0.016) derived from in silico metagenomics. Samples were grouped in > 75th percentile relative abundance and ≤ 75^th^ percentile relative abundance. **F** non-metric multidimensional scaling plot based on relative abundance of 17 relevant functional pathways for glycocalyx degradation predicted by PICRUSt-2 (nMDS, Bray-Curtis Similarity, square root data transformation, stress value: 0.09), each dot represents one sample, colour indicates corresponding BT, vector length and direction indicates impact of the respective pathway on sample distribution within the plot.

**Table 1.** Sialidase encoding genes identified in bile samples through shot gun metagenomic analysis. The encoded proteins and the RKPM values mapping to the genes are given.

| gene | sample ID | 025 | 082 | 095 | 100 | 115 | 127 | 128 | 135 | 155 | 158 | 174 | 182 | 217 | 218 |
| --- | --- | --- | --- | --- | --- | --- | --- | --- | --- | --- | --- | --- | --- | --- | --- |
| Q8XMY5 Clostridium perfringens 13 NanJ VAL |  |  |  |  | 0.38 | 3.49 | 0.84 |  | 0.22 |  | 0.49 |  |  | 2.26 | 19.80 |
| P10481 Clostridium perfringenc A99 NanH VAL |  |  | 0.72 |  |  | 2.07 | 0.64 |  |  |  |  |  |  | 2.31 | 15.15 |
| WP_011590331 Clostridium perfringens ATCC13124_NanI VAL |  |  | 0.60 |  |  | 4.92 | 0.96 |  | 0.38 | 0.51 |  |  |  | 1.97 | 16.61 |
| P77848 Clostridium tertium DSM2485 NanH Val |  |  |  |  |  | 0.55 |  |  |  |  |  |  | 1.06 |  |  |
| WP_003456009 Clostridium perfringens ATCC13124 |  |  |  |  |  | 2.14 |  |  |  |  |  |  |  | 1.83 | 14.04 |
| EFM35933 Streptococcus sp. 73H25AP |  |  | 24.96 |  |  | 0.31 |  | 0.36 | 33.08 |  | 0.49 |  |  |  |  |
| EFO55109 Streptococcus infantis SK1302 |  |  |  |  |  |  |  |  |  | 1.67 |  |  |  |  |  |
| KXU00741 Streptococcus oralis DD30 |  |  |  |  |  |  |  |  | 36.38 |  |  |  |  |  |  |
| EFE56037 Streptococcus oralis ATCC_35037 |  |  | 4.53 |  |  |  |  |  | 16.70 |  |  |  |  |  |  |
| P62576 Streptococcus pneumoniae ATCC BAA255 NanA VAL |  |  | 0.80 |  |  |  |  |  | 1.86 |  |  |  |  |  |  |
| Q54727 Streptococcus pneumoniae ATCC BAA334 NanB VAL |  |  |  | 6.37 |  |  |  |  |  |  |  |  |  |  |  |
| WP_009406364 Actinomyces oris ERR9969204 |  |  |  | 3.97 |  | 0.40 |  |  |  |  | 0.38 |  |  |  |  |
| WP_060956757 Actinomyces oris T14V NanH VAL |  |  |  | 2.91 |  |  |  |  |  |  | 0.21 |  |  |  |  |
| WP_075411946 Actinomyces oris R2109 |  |  |  | 1.82 |  |  |  | 0.30 |  |  | 0.36 |  |  |  | 0.11 |
| WP_009405803 Actinomyces sp. F0384 |  |  |  | 2.49 |  |  |  | 0.63 |  |  | 0.42 |  |  |  |  |
| WP_009747907 Actinomyces_sp. F0384 |  |  |  | 2.92 |  |  |  |  |  |  |  |  |  |  | 0.08 |
| WP_003790542 Schaalia odontolytica ATCC17982 |  |  |  | 0.35 |  |  |  |  |  |  | 2.34 | 2.40 |  | 1.29 |  |
| WP_003792757 Schaalia odontolytica ATCC17982 |  |  |  |  |  |  | 0.09 |  |  |  | 1.18 | 0.73 |  | 1.41 |  |
| WP_003828032 Sial Bifidobacterium breve DSM20213 |  |  |  |  |  |  |  |  |  |  |  |  |  | 6.42 |  |
| WP_311371633 Bifidobacterium dentium |  |  |  |  |  |  |  |  |  |  | 1.02 |  |  |  |  |
| A6L7T1 Phocaeicola vulgatus ATCC8482 NanH VAL |  |  | 1.19 |  |  |  |  |  |  |  |  |  |  |  |  |
| WP_007135677 Prevotella salivae DSM15606 |  |  |  |  |  |  | 0.08 |  |  |  | 1.21 |  |  | 0.43 |  |
| WP_010755296 Enterococcus pallens BAA351 |  | 1.64 |  |  |  |  |  |  |  |  |  |  |  |  |  |
| WP_049016767 Klebsiella pneumoniae W14 |  |  |  |  |  | 0.31 | 3.96 | 1.06 | 6.31 |  |  |  |  |  |  |
| WP_276855804 Enterocloster lavalensis |  |  |  |  | 1.89 |  |  |  |  |  |  |  |  |  |  |
| WP_147735841 Brachyspira aalborgi PC5538III-Ic |  |  |  |  |  |  |  |  | 1.11 |  |  |  |  |  |  |
| CDA74708 Bacteroides sp. CAG:530 |  |  |  |  | 1.38 |  |  |  |  |  |  |  |  |  |  |
| MBP3481490 Clostridia bacterium |  |  |  |  |  |  |  |  | 1.75 |  |  |  |  |  |  |

Genes are given as accession number, bacterial strain and (if needed) name of sialidase. Three digit number in column header displays sample ID. RKPM = reads per kilo base per million mapped reads

### Bacterial communities influence course of primary sclerosing cholangitis

Groups of samples dominated by *Enterococcus, Streptococcus, Staphylococcus,* Enterobacteriaceae, or not dominated by any genus (= 5 groups) were further evaluated. In cross-sectional analysis, BTs could not be correlated to the clinical-morphological type of PSC and co-occurrence with either inflammatory bowel diseases (Extended Data Table 2). Laboratory parameters were similar in all groups except for alkaline phosphatase and gamma glutamyltransferase with higher values in *Enterococcus* BT compared to no BT in the post-hoc analysis.

We then analyzed whether the presence of BTs in bile of patients correlated with a poor or good disease prognosis. It is well accepted that parameters measured in blood do not necessarily reflect local biliary inflammatory activity in PSC patients^11^. Therefore, we used relevant clinical endpoints such as the need of liver transplantation, development of cholangiocellular adenocarcinoma (CCA) and death for an event-free survival analysis. Patients with no BT had the best event-free survival, patients with an Enterobacteriaceae or *Enterococcus* BT an intermediate and patients with a *Streptococcus* or *Staphylococcus* BT the worst event-free survival (no BT vs. *Streptococcus* BT p = 5.9×10^-4^, no BT vs. *Staphylococcus* BT p = 0.009, *Streptococcus* BT vs. Enterobacteriaceae BT p = 0.006, Figure 1B).

### Potential for glycocalyx degradation explains different long-term outcome

Next, we tried to identify bacterial pathogenic functions that may drive disease activity. Different bacterial species can share pathogenic functions leading to a similar microbiome-host-interaction despite taxonomic differences^12,13^. Since *Streptococcus* BT and *Staphylococcus* BT were correlating with a poor event-free survival, we hypothesized that those two BTs were associated to a disadvantageous functional community structure. Therefore, samples were regrouped according to the presence of such a disadvantageous BT in the history of the donating patient. Based on this new binary grouping, event-free survival analysis revealed a pronounced superiority of patients without a disadvantageous BT in their history (p = 0.001, Figure 1C).

In order to identify pathogenic functions possibly responsible for the disadvantageous character of microbial communities, we applied a machine learning approach on all functional pathways predicted by *in silico* metagenomics (PICRUSt-2^14^, output of MetaCyc pathways^15^ output) comparing samples from patients without vs. with a disadvantageous BT in their history. From 392 pathways, three pathways involved in the degradation of glycocalyx components were among the results top five pathways differentiating best between the two groups (Extended Data Table 3). Pairwise comparison revealed the superpathway of *N*-acetylneuraminate degradation (GLCMANNANAUT), comprising enzymes involved in the degradation of sialic acid (encoded by the *nan*-gene cluster), as well as the pathway for lactose and galactose degradation I (LACTOSECAT) being higher abundant in the group of samples with a disadvantageous BT in patient history. In order to verify the clinical relevance of those findings, Event-free survival analysis of samples grouped according to their relative abundance of those two pathways in either >75^th^ percentile or ≤75^th^ percentile revealed a significantly reduced event-free survival of patients with a high relative abundance of either pathway (GLCMANNANAUT: p = 0.006, Figure 1D; LACTOSECAT: p = 0.011, Figure 1E). Focusing on those functional pathways involved in the degradation of known cholangiocyte glycocalyx components (n = 17, Extended Data Table 4), the multidimensional scaling plot visualized similar patterns in Streptococcus and Staphylococcus BTs with GLCMANNANAUT and LACTOSECAT as main pathways differentiating them from samples with other BTs (Figure 1F). Thus, results from the *in silico* approach suggested bacterial degradation of cholangiocyte glycocalyx components as relevant pathogenic functions in patients with severe courses of PSC.

### Confirmed presence of sialidase genes and active sialidase in human bile

The glycocalyx of eukaryotic cells is a dense layer of membrane bound peptidoglycans. Sialic acid is located in the terminal position of cholangiocyte glycocalyx^3^. This localization is crucial for cell protection against cytotoxic bile acids^4^. Thus, sialic acid is directly exposed to bile and is easily accessible to bacteria of the biliary microbiome. While enzymes or transporters involved in the bacterial degradation of sialic acid are included in the GLCMANNANAUT superpathway, sialidase, the first enzyme needed to cleave off sialic acid from host glycoconjugates, is not included. Therefore, the relative abundance of sialidase (EC 3.2.1.18) predicted by the same *in silico* metagenomics analysis was used additionally. Patients with a predicted relative abundance of sialidase <25^th^ percentile were compared with patients with a relative abundance >75^th^ percentile. The result confirmed an inferior event free survival in patients with a high predicted relative abundance of sialidase (Figure 2A).

**Figure 2:**
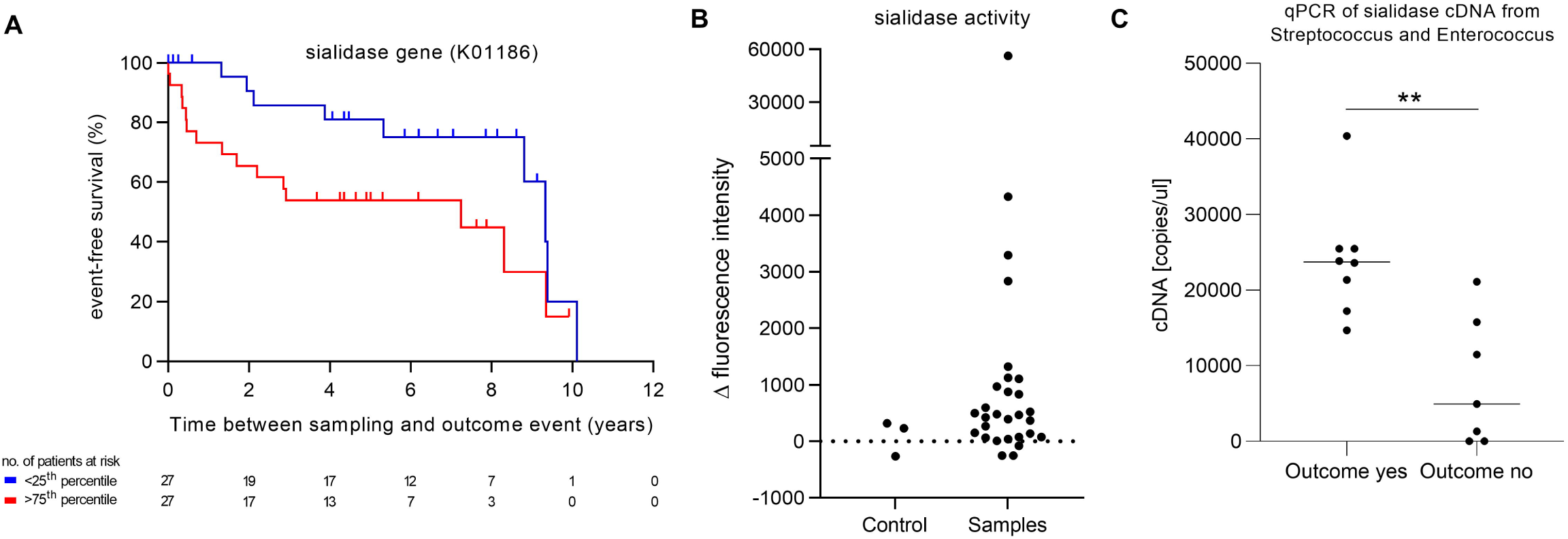
Correlation of clinical course of PSC with the presence of bacterial sialidase genes and mRNA and measured sialidase activity in bile. **A** The event-free survival of PSC patients according to relative abundance of Sialidase-1 (EC:3.2.1.18) derived from *in silico* metagenomics. Outcome defining events: liver transplantation, cholangiocellular adenocarcinoma (CCA) and death. Samples with a relative abundance > 75th percentile and < 25th percentile were included and compared. p = 0.049 **B** Fluorimetric Sialidase activity assay with 4-MU-Sia as a fluorogenic substrate. ΔFluorescence intensity (= Fluorescence intensity (60 min) – Fluorescence intensity (0 min)) is shown. Exemplarily, heat inactivated bile samples served as negative control. **C** quantitative PCR of sialidase mRNA (cDNA) from Streptococcus and Enterococcus. Outcome yes: n = 8, Outcome no: n = 7, p = 0.002, bar indicates median.

We established a quantitative fluorometric sialidase activity assay with 2’-(4-Methylumbelliferyl)-Sia as a substrate. Enzymatic removal of sialic acid led to the production of fluorogenic 4-methylumbelliferone (4-MU). As expected from variable clinical courses, sialidase activity varied between bile samples (Figure 2B). Heat inactivated samples from PSC patients were used as a control group. Due to the scarcity of samples, only a few were heat inactivated. Those samples with a high sialidase activity were donated by patients with a severe clinical course in young life, underlining a potential role of sialidase for the clinical course of patients with PSC. Mass spectrometry-based analysis of two bile samples of PSC patients resulted in the identification of 1059 human proteins. Human sialidases could not be identified (Supplementary Information 1).

Next, we aspired the identification of bacterial species that are able to produce a sialidase in bile. First, a search for validated sialidases encoded in published complete genomes or draft genomes of bacterial species prevalent in bile was conducted (Supplementary Information 2, Extended Data Figure 3). The results revealed sialidase genes in the core genome of *Streptococcus pneumoniae* or *infantis* and of high amounts of *S. mitis* or *S. oralis* strains as well as in the core genome of *Clostridium perfringens, Actinomyces naeslundii/viscosus/oris, Bacteroides fragilis* and *Cutibacterium acnes* (Supplementary Information 3). *Enterococcus* species, except *E. avium* and *E. durans*, only seldomly (E. *faecalis, E. casseliflavius)* or never *(E. faecium)* encode a sialidase. Interestingly, in *Staphylococcus spp.* The presence of a sialidase is restricted to *S. schleiferi, S. pseudointermedius* and *S. delphini*, species which were not observed in bile samples (Supplementary Information 3).

Next, we performed shot gun metagenomics in 20 bile samples. The results confirmed the presence of sialidase genes in bile samples with high abundances of *Clostridium perfringens, Actinomyces/Schaalia* or organisms related to *Streptococcus oralis* and *infantis*. (Table 1, Extended Data Figure 3). As expected from phylogenetic analysis all three NanH, I and J encoding genes were identified in *C. perfingens* comprising samples in similar abundances. Sialidases encoded by *Bifidobacterium* or *Enterococcus pallens* were identified as expected. *Enterococcus spp*. and *Streptococcus spp*. are of major interest in terms of the clinical course in PSC patients. Therefore, bacterial species were isolated from bile and their genomes sequenced. Only few *Enterococcus faecalis* and no *E. faecium* isolate so far are reported to encode sialidase or a *nan*-gene-cluster. We isolated and sequenced 100 *Enterococcus strains* from bile to identify possible genomic adaptations in this niche (including 45 *E. faecalis* and 36 *E. faecium* plus 9 related *E. lactis*) (Supplementary Information 4). However, only one of these 90 *E. faecalis/faecium/lactis* isolates – namely strain FR191 – revealed the presence of a sialidase gene and the whole *nan*-gene-cluster together on one operon. Besides, only one *E. galllinarum* and the *E. maldoratus* isolate obviously encode a sialidase (Extended Data Table 5).

So far reported, *Streptococcus* strains related to S*. infantis, S. mitis, S. pneumoniae, S. oralis* and others, which are difficult to be distinguish by amplicon sequencing, may encode different sets of sialidases (Supplementary Information 3). We isolated seven *Streptococcus* strains related to *S. mitis, S. oralis* or *S. infantis* from bile (Extended Data Figure 4). Strikingly, genome sequencing indicated at least one sialidase encoding gene in all seven isolates related to those species (Extended Data Table 5, Extended Data Figure 3).

Continuatively, to analyze if those genes are expressed in bile, sets of primer for an mRNA based qPCR approach identifying sialidase sequences of sialidases from Streptococcus and Enterococcus were designed based on identified sialidase sequences in the phylogenetic analysis. Fifteen randomly picked samples from different patients were analyzed. Bacterial sialidase mRNA could be detected in 13 of those samples. In line with findings from *in silico* analysis, the concentration of mRNA was higher in samples derived from patients with a severe clinical outcome (with outcome: n = 8, without outcome: n = 7, p = 0.002, Figure 2C).

Thus, in bile, sialidases can be produced by different species across the phylogenetic tree, the amount of that production can be quantified and correlates with the clinical outcome of individual patients.

### Desialylation and bile acid exposure to cholangiocytes induces apoptosis

In order to analyze the impact of bacterial sialidases on cholangiocyte biology *in vitro*, the sialidase from *Enterococcus faecalis* FR191 was further investigated. For this, in the supernatant of *Enterococcus faecalis* FR191 (E.f.), grown for 48 h in complex medium, sialidase activity was detected in stationary growth phase and the activity was enhanced when growth proceeded in the presence of sialic acid (mean delta absorbance ± standard deviation 1.14 ± 0.11 vs. 2.26 ± 1.3; n = 4; p = 0.029). This enhancement appeared to be highest during mid-stationary phase. To assign the sialidase activity to the gene identified by sequence homology, the E.f. sialidase gene was subsequently cloned, recombinantly expressed in *E. coli* and purified by an affinity chromatography for use in cell culture experiments. In a commercially available and immortalized cholangiocyte cell line MMNK-1 the presence of sialic acid in terminal position of the cholangiocyte glycocalyx was confirmed by lectin immunofluorescence stainings with *Maackia amurensis* lectin (MAL-II) and *Sambucus nigra* agglutinin (SNA) detecting α2,3-and α2,6-linked sialic acid, respectively. Enzymatic removal of sialic acid by treatment of MMNK-1 cells with the purified E.f. sialidase lead to the appearance of a positive staining with the Peanut agglutinin lectin (PNA), detecting terminal galactose (Figure 3A), thus proving the removal of sialic acid and the identification of galactose as the underlying monosaccharide in the glycan. To further validate the ability of different bacterial sialidases to desialylate cholangiocytes, two sialidases from two different *Streptococcus mitis* strains were further investigated as well. Both sialidase genes were found in biliary bacteria. The sequences were also subsequently cloned, recombinantly expressed in *E. coli* and purified by an affinity chromatography for use in cell culture experiments with the MMNK-1 cells. The ability to desialylate the cells was proven In FACS analysis (Figure 3D).

**Figure 3:**
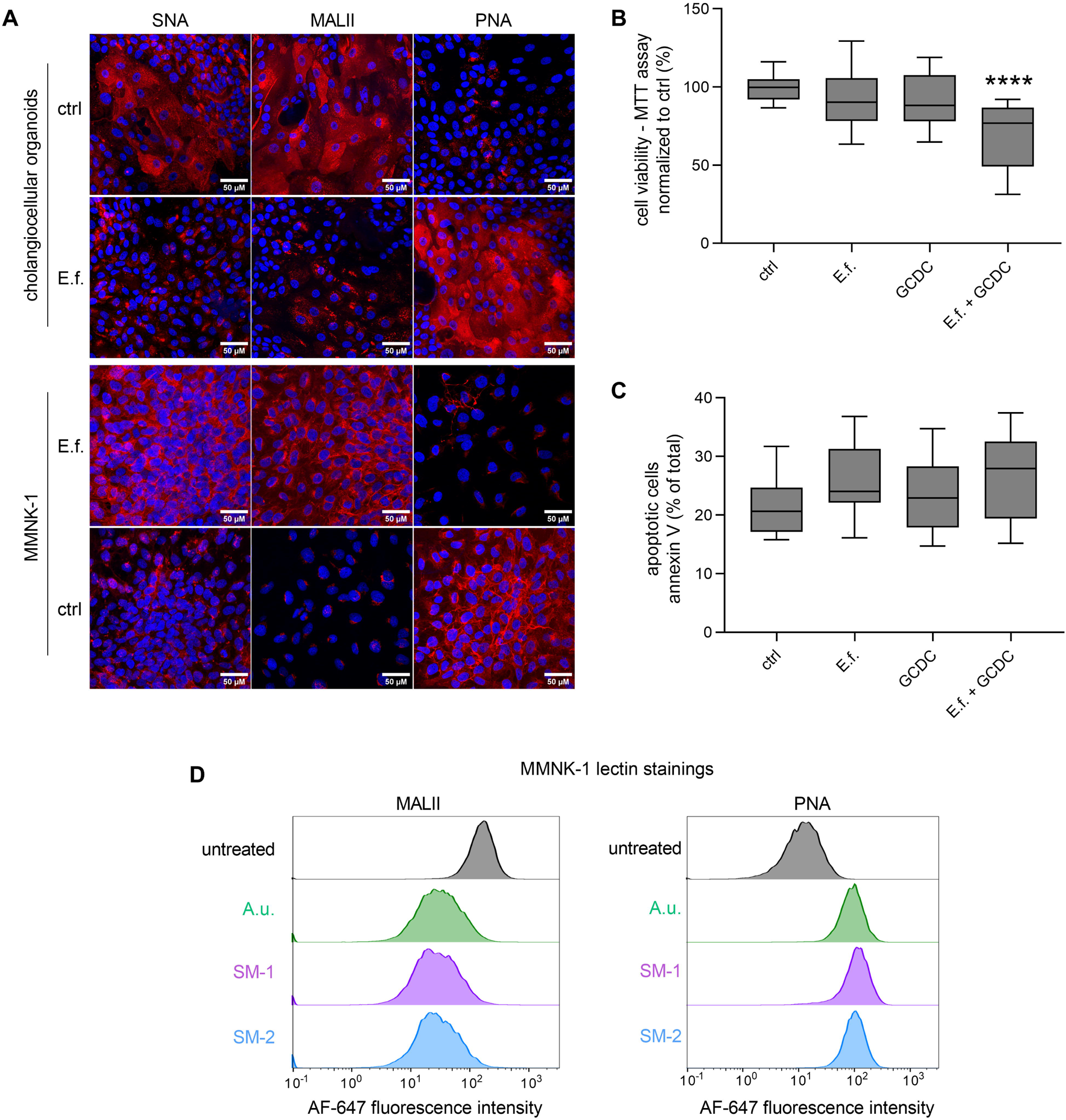
Desialylation enhances bile acid induced toxicity in 2D and 3D cholangiocyte cultures. **A** Representative images of immunofluorescence stainings of cholangiocellular organoids and MMNK-1 cells. Stainings with DAPI (blue) and biotinylated lectins SNA (staining α2,6 linked sialic acid), MALII (staining α2,3 linked sialic acid) and PNA (staining β1,3 linked galactose) (red). Scale bars, 50 µM, nt = non treated **B** Viability of cholangiocellular organoids after treatment with Enterococcus faecalis (E.f.) sialidase, bile acid (GCDC) or treatment with GCDC after desialylation with E.f. sialidase. Assessed by the MTT assay. Experiments (n = 6) were performed in triplicate and data are shown as boxplots with mean ± standard error of the mean. **** p < 0.0001 one-way ANOVA, and post-hoc Dunnett’s multiple comparison tests. **C** Annexin V staining of cholangiocellular organoids analyzed via flow cytometry. Cells were treated with either E.f. sialidase, GCDC (bile acid) or a combination of both. Experiments (n = 4) were performed in triplicate and data are shown as boxplots with mean ± standard error of the mean. **D** Flow cytometrie histograms of the lectin staining of MMNK-1 cholangiocytes: live cells stained with lectins MALII (staining α2,3 linked sialic acid) and PNA (staining β1,3 linked galactose) with or without treatment with Streptococcus mitis sialidase A or B.

Host-genetic factors play a relevant role in the pathogenesis and disease severity of PSC^16,17^. In order to exclude this factor and still respect individual susceptibilities we generated patient derived cholangiocellular organoids via cytology brushes or from fresh bile samples of control patients and utilized them in cell culture experiments. Different lectin immunofluorescence staining confirmed the architecture of the glycocalyx in patient derived cholangiocytes with α2,3-and α2,6-linked sialic acid in the terminal position indicated by SNA and MAL-II staining and β1,3-linked galactose as the underlying sugar molecule visible after E.f. sialidase treatment (Figure 3A).

To analyze the impact of desialylation of cholangiocytes in the biliary system of patients, the physiological biliary environment was modeled by treating cholangiocyte organoids with the bile acid glycochenodeoxycholic acid (GCDC). Viability assays revealed a significantly higher susceptibility for bile acid induced damage after desialylation with bacterial sialidase compared to sialidase or GCDC treatment alone (Figure 3B). Apoptosis assays showed a similar tendency (Figure 3C). Thus, bacterial sialidases in bile can change the cholangiocyte glycocalyx architecture leading to a higher susceptibility towards cytotoxic effects of bile acids promoting disease progression. Next, we investigated cholangiocyte sialylation in liver biopsies of PSC patients by immunofluorescent lectin analysis and observed an inhomogeneous distribution of sialic acid between different samples and within each sample (Extended Data Figure 2). Thus, impairment of the cholangiocyte protective shield is varying between different patients and additionally along the biliary tract of individual patients. This inhomogeneous disease pattern correlates with the known variable courses of PSC in clinical observations as well as with the asymmetric morphological disruption of the biliary tract in PSC patients.

## Discussion

With this study we identified a new factor in the pathogenesis of PSC. Members of the biliary microbiome initiate degradation of the cholangiocellular glycocalyx by sialidases. In consequence, the protective shield of cholangiocytes against toxic bile acids is destroyed leading to an increased susceptibility to bile acid toxicity which is known to drive inflammation and destruction of the biliary tract in inflammatory bile duct associated diseases like PSC^2,8,18^. A difference in the long-term outcome of PSC could be demonstrated based on sialidase activity and sialidase gene transcription according to the observations from *in silico* metagenomics.

Some recent interventional studies have indicated a participation of bacteria in the pathogenesis of PSC as antibiotic treatment or fecal microbiome transfer attenuated disease activity^9,19^. However, the results were incongruent with some patients benefiting from the interventions while others did not. Our study now substantiates the involvement of bacteria on PSC by identifying patterns in the microbiome that correlate with a poor prognosis of affected patients. Bacterial glycan degrading capabilities were identified as crucial pathogenic element. Nutrients for bacteria in bile are rare especially in cholestatic diseases with impaired bile flow. Thus, utilizing glycocalyx components as energy source most likely displays a survival advantage in that niche. Different bacterial species present in the biliary microbiome have the capability to degrade sialic acid as terminal saccharide in the cholangiocytes glycocalyx. Once sialic acid is cleaved off, other saccharide compounds are exposed to bile and might also be utilized by other members of the bacterial community as energy source. With degradation of the glycocalyx and especially the loss of sialic acid and by that the loss of the negative charge, cholangiocytes lose the effective protection against cytotoxic bile acids, similar to results reported previously^4^.

Thus, different bacterial species contribute to the pathologic functional microbiome pattern. Those findings strongly support the idea of pathogenic functions as virulence factors and the ides of adding functional dysbiosis of the microbiome to the classical definition of infections^20^. Additional analyses including transcriptomics and a validation of the functional dysbiosis as predictive marker for clinical course of disease in prospective clinical studies will be needed for further evaluation. In this study hard clinical endpoint were used in order to identify patterns sufficiently within high dimensional and individually highly variable data sets. Analyses in further prospective cohorts should regard graduated disease severity, i.e. Mayo PSC risk score, as well.

In order to do so, standard clinical microbial diagnostics should exceed taxonomic analysis and include analysis of functional properties. As standard microbial cultivation of bile identifies only some but not all members of the microbial community^21^ sequencing techniques will be needed for a proper analysis. A targeted transcriptomic approach analyzing bacterial mRNA of sialidase genes and genes coding for enzymes involved in the degradation of galactose and sialic acid in addition to taxonomic profiling and quantification of the respective enzymes in bile could be a valuable and practical approach. For this approach, obtaining bile via endoscopic retrograde cholangiography (ERC) is a requirement. So far clinical diagnostic concentrates on bile duct morphology and blood parameters. Due to the safety and progressively increasing image quality of magnet resonance imaging (MRI) based cholangiography, the role of ERC in the diagnostics of bile duct associated diseases diminished over the past decade and indication is concentrated on interventional approaches^22,23^. Together with recent findings that blood parameters do not properly correlate with inflammatory processes in bile^11^, this study supports the necessity of diagnostic ERCs. The risk of severe complications of that procedure have, however, to be evaluated properly. If morphological analysis is done via MRI, diagnostic ERC could focus on obtaining bile without papillotomy or application of contrast agents for fluoroscopy, which will substantially reduce the risk for severe complications of a diagnostic ERC.

For evaluation of pathological microbiome-host-interactions the host site, in particular the individual architecture of the cholangiocyte glycocalyx, has to be analyzed in clinical diagnostic as well. The International PSC Study Group recently identified mutations in genes involved in the assembly of glycocalyx peptidoglycans like FUT-2 in PSC patients^16,17^. Thus, the resilience towards bacterial degradation of cholangiocytes protective shield most likely varies between individual patients. We assume that dysbiosis alone does not lead to PSC without the co-occurrence of a vulnerable host glycocalyx. So far, little is known about the specific architecture of cholangiocytes glycocalyx both, *in vitro* and *in vivo*. Further studies are needed to evaluate the impact of that structure on bile duct homeostasis in health and disease. Organoids are a suitable model for this evaluation since they can be generated from cytology brushes of individual patients allowing an individualized diagnostic.

Knowing the functional patterns of the biliary microbiome and the individual cholangiocytes resilience towards glycocalyx degradation may help to identify patients at risk for a severe clinical event like the need of liver transplantation, development of CCA or death. By resolving the dysbiosis and inhibiting the bacterial degradation of the cholangiocyte glycocalyx, the course of disease could be modified. So far, no disease modifying treatments are available for PSC. Treatment options are limited to management of complications like bile duct obstructions via ERC and/or bacterial cholangitis by administration of antibiotic medication^1^. Findings from this study could open new targeted treatment strategies based on functional patterns in the individual biliary microbial community. Further studies will be needed to evaluate the potential use and harm of such therapeutic approaches.

With those findings, our study changes the clinical view on pathologies in the bile duct. This study prepares a basis for implementing biomarkers in the clinical diagnostic that might allow to identify patients at risk for a severe clinical event and poor prognosis. Bacterial functional dysbiosis is identified as crucial pathogenetic and disease modifying factor. Redefining PSC as a partially infectious disease would widen diagnostic and disease modifying therapeutic approaches allowing to breach the helplessness affected patients and attending physicians are currently facing with this disease.

## Material and Methods

### Patient population

All bile samples were derived from patients with PSC via ERC during in-patient treatment at Hannover Medical School between 2008 and 2014 and were directly stored at-80 °C. Clinical data and laboratory results were obtained retrospectively from electronic health records. In total 209 bile samples from 117 different patients with PSC were included. Forty-four patients donated more than one sample (maximum: 10 samples). Samples of the validation cohort (n = 56) were sampled and stored between 2017 and 2022 accordingly.

### Sample preparation, sequencing and bioinformatical pipeline

DNA extraction and amplicon preparation were done as described previously^24^. Sequencing data of the validation cohort were processed via the Divisive Amplicon Denoising Algorithm (DADA2)^25^. The ribosomal database project (RDP) naive Bayesian classifier was used to classify the phylotypes by applying a confidence threshold of 80 %^26^. Within each sample, sequence counts were merged at taxonomic levels. Relative abundance of one taxon was defined as the percentage of reads for this taxon from total read count of the respective sample. *In silico* metagenomics were done using PICRUSt-2 (input: biome-file with reference sequences. MetaCyc database^15^ and KEGG output was further used. Date January 2021)^14^.

### Metagenomics and Genomics

DNA libraries of bacterial isolates were created using the NEBNext Ultra II FS DNA Library Preparation Kit. The libraries were checked with an Agilent technology Bioanalyzer 2100. Paired-end libraries were sequenced using Illumina v3 chemistry on an Illumina MiSeq sequencer with a 250-bp paired-end protocol. The qualified sequence reads were assembled by *de novo* assembly using Unicycler^27^, an assembly pipeline provided by Genome Assembly Service of BV-BRC (Bacterial and Viral Bioinformatics Resource Center, https://www.bv-brc.org). The assembly quality was evaluated by comparison to corresponding complete reference genomes using QUAST^28^. After assembly, the draft contigs were annotated using the BV-BRC Genome Annotation Pipeline, which uses the RAST tool kit (RASTtk) to provide genomic features and predicted genes^29^. The TYGS server was used to query by whole genome comparison if strains represent novel species or belong to known species^30,31^. Digital DDH values and confidence intervals were calculated using the recommended settings of the GGDC 4.0. For the phylogenomic inference, all pairwise comparisons among the set of genomes were conducted using GBDP and accurate intergenomic distances inferred under the algorithm ‘trimming’ and distance formula d5^32^. If strains probably represent novel species, the closest related type strain is given (Supplementary Information 4) as well as other related species (Extended Data Figure 5)

Forty-four sialidases of validated function as well as 19 probable sialidases (Supplementary Information 2) were used to identify sialidases encoded in genomes of novel isolates. These sequences were also used as seeds to characterize the spread of respective genes in bacterial taxa from bile (Supplementary Information 3). The number of genomes searchable for sialidase encoding genes was defined by the number of genomes encoding rpoD. Both the NCBI genome database (tblastn, used against all complete genomes as well as all draft genomes of a given taxon) and the BV-BRC database (https://www.bv-brc.org/app/Homology) were used.

Twenty shot gun metagenomic sequence datasets were produced from total DNA extracts of bile samples obtained with the MOBIO PowerSoil DNA Isolation Kit (QIAGEN) according to the manufacturer’s instructions. Libraries were prepared using NEBNext Ultra II FS DNA Library Prep Kit without fragment size selection and paired-end libraries were sequenced on a NovaSeq 6000 SP with the Reagent Kit v1.5 (Illumina, 300 cycles). Demultiplexed raw reads were trimmed for low quality and filtered against the phix174 and human hg19 genome. The remaining 122.143.898 reads (20 datasets with a minimum of 1.977.008 and a maximum of 24.367.144 reads) were analyzed for reads associated with sialidase encoding genes.

A sialidase dataset was built using genes encoding proteins belonging to the KEGG orthologous group K01186 (EC 3.2.1.18 exo-alpha-sialidase) from the 9.9 million gene integrated reference catalogue of the human microbiome (IGC)^33^ supplemented with gene sequences encoding 44 validated and 19 putative sialidases (listed in Supplementary Information 2) and genes of the IGC, the encoded proteins of which showed a sequence identity >50 % to any of the proteins listed in Supplementary Information 2. All quality filtered libraries were mapped against this sialidase dataset comprising 1086 unique gene sequences (Supplementary Information 5) using BBMap Version 38.84 (Supplementary Information 6).

To compare gene coverage values the RPKM (Reads per kilo base per million mapped reads) was calculated which corrects differences in both: sample sequencing depth and gene length. Only genes which accumulated >1 RPKM and ≥5 reads in at least one sample were considered and a representative protein with >99 % identity was defined. Finally reads for all genes where the encoded proteins shared >90 % identity were summed up (Table 1).

### *In silico* prediction of disadvantageous community structure

A linear regression model with elastic net regularization was trained on pathway abundances to predict whether individuals had a history of disadvantageous community structure.

Pathway abundances that did not vary across samples were removed (7 out of 392 pathways). Missing values were imputed using median imputation (46 measurements, 0.05 % of the total dataset). Values were rank-normalized prior to further analysis.

The model was trained using a five-fold repeated cross-validation design on 70 % of the data, and the final model’s performance was evaluated on the remaining, unseen 30 % of the data using the R package caret^34^. Hyperparameters alpha and lambda were constrained to [0.01, 0.09] and [0, 1], respectively. To account for class imbalance, weights were given to each class proportional to their prevalence. Model optimization was performed based on the Area under receiver-operator characteristics (AUC) metric. To assess the reproducibility of these results, we performed one hundred independent iterations of model training and performance assessment and report the median AUC across these iterations. To interpret the obtained results, we assessed the average coefficient per pathway as well as how frequent each pathway was included in the final model.

### Fluorimetric Sialidase activity Assay

Bile samples have been stored at-80°C. Sialidase activity was measured in 10 µl of thawed bile samples using 1 µM f.c. 4-Methylumbelliferyl)-a-D-*N*-acetylneuraminic acid (4MU-Sia, Biosynth, EM05195) in 30 µl 50 mM sodium acetate buffer pH 5.5 before and after incubation at 37°C for 60 min as previously described^35^. Samples were measured in three technical replicates with a BioTek Synergy Mx Microplate Reader (excitation at 365 nm and emission at 448 nm) after adding 100 µl of the the stop solution (0.2 M glycine buffer, pH 10.7). Δ Fluorescence intensity” = Fluorescence intensity (60 min) – Fluorescence intensity (0 min) was calculated. Heat-inactivated samples of PSC patients were used as a control group.

### Mass-Spectrometry

Two bile samples were protected by adding protease inhibitor cocktail (1:100, Sigma) followed by centrifugation at 13,000 g and 4 °C for 10 min. The supernatant was mixed with 4 volumes of ice-cold 0.1 M ammonium-acetate in methanol and incubated for 16 hours at-20 °C. After centrifugation for 15 min at 13,000 g and 4 °C, the supernatant was discarded and the pellet was washed twice with ice-cold 0.8 M ammonium acetate in 80 % methanol and twice with ice-cold 80 % acetone, with centrifugation steps for 15 min at 13,000 g and 4 °C in between followed by drying the protein pellet at 20 °C after the last washing step. Pellets were re-suspended in 36 µL 1x Laemmli buffer, incubated at 95 °C for 5 min and further purified by SDS-PAGE with a 4 % stacking gel and a 12 % resolving gel. Protein-containing SDS gels were stained with Coomassie (Biorad) over night and destained with destaining solution (Biorad) for 2 h. Each lane was cut into 8 fractions restricted to proteins > 20 kDa. Each fraction was further chopped followed by in-gel digestion with Trypsin Gold (Promega) following manufacturer’s instructions.

Peptides were resuspended in 10 µL water with 5 % acentonitrile (ACN) and 0.1 % formic acid (FA) and total sample was analysed via liquid chromatography–mass spectrometry using a Vanquish Neo nanoflow UHPLC System coupled to an Orbitrap Eclipse mass spectrometer (Thermo Scientific™). Peptides were separated on a reversed-phase nanoViper™ PepMap™ separating column (150 mm length, 75 μm inner diameter, and 2 μm C18 particle size (Thermo Scientific™)) using buffer A (80 % ACN, 0.1 % FA) and B (80 % ACN, 0.1 % FA). Peptides were eluted at a flow rate of 250 nL/min at 40 °C by a gradient starting with Buffer B increased from 6 % to 25 % within 60 min and further increased to 90 % within 10 min and hold at 90 % for additional 8 min. Ionisation was achieved by a Nano Spray Flex Ion Source and stainless steel emitters (40mm, OD 1/32) at 1,900 V. Precurser and fragment ions were both scanned in the Orbitrap analyser. Precurser scans were performed with an m/z range set to 375-1500 and with a resolution of 120,000, stored in profile mode. The twenty most intense precursors with intensities above 2,000 counts were fragmented in the linear ion trap by higher-energy collisional dissociation with collision energy set to 30 % and dynamic exclusion set to 45 s. Fragment ion scans were performed at a resolution of 15,000, with a normal mass range and were stored in centroid mode.

MS raw data were processed with MaxQuant software (V. 2.2.0.0)^36^ for the identification and quantification of proteins using preconfigured settings except for label-free quantification which was set to IBAQ. All raw files derived from the same sample were grouped into separate experiments with multiple fractions. MS spectra were searched against UniProt databases of human proteins (reviewed, database downloaded on 16/08/2023) and all known sialidases and neuraminidases (reviewed, database downloaded on 03/11/2023). The resulting list of protein groups was filtered for proteins not classified as contaminants, proteins identified in the reverse database or only identified by site modifications and further screened for proteins with protein names containing strings *sialidase* or *neuraminidase* (Supplementary Information 1).

### Isolation of bacteria from bile

Native bile samples were immediately transferred to the laboratory in sterile tubes or syringes and cultured in accordance with DIN EN ISO 15189 and common clinical microbiology methodology^37^. Briefly, chocolate agar (GC II Agar with IsoVitaleX, BD), sheep blood agar (BBL Columbia Agar, BD), MacConkey agar (Mast Diagnostica, Reinfeld, Germany or BD), tryptone soy broth (Oxoid or Xebios), schaedler agar (BD), schaedler agar supplemented with kanamycin and vancomycin (Schaedler KV Agar, BD), anaerobe basal broth (Oxoid), mannitol salt agar (BD) and mal extract agar (Xebios) were inoculated with native bile. Chocolate agar was incubated for 48h at 37°C in a 5 % CO_2_ atmosphere, schaedler agar, schaedler KV agar and anaerobe basal broth were incubated for 7d at 37°C in an anaerobic atmosphere and other media were incubated for 48h at 37°C in normal atmosphere. Media were observed for growth after at least 18h of incubation and bacteria were subsequently identified using MALDI-ToF (Vitek MS, Biomerieux) and isolated on sheep blood agar. Isolated bacteria were frozen in tryptone soy broth supplemented with 30 % glycerin at-80°C until DNA extraction and whole genome sequencing.

### Cell Culture

MMNK1 Cells (JCRB1554) were cultured in DMEM F12 (Gibco) supplemented with 1 % Penicillin-Streptomycin and with or without 5 % FCS. Starving medium was prepared as described above, without the addition of FCS. Cells were kept at 37°C in a 5 % CO_2_ atmosphere and passaged twice a week. Cholangiocellular organoids were cultured embedded in extracellular matrix solution (90 % MatrigelTM, 10 % expansion media) and the extracellular matrix dome was overlayed with expansion media (Extended Data Table 6). Organoid cultures were kept at 37 °C and 5 % CO_2_ and passaged once per week with TrypLE ExpressTM (Gibco).

### Spectrophotometric sialidase activity assay

Enterococcus spp. FR191 and FR143 were cultivated for 48 h in 12.5 ml of Müller-Hinton-Infusion medium with or without 22.3 µM sialic acid (pH 7.3). Bacteria were incubated at 37 °C for 48 h. Growth was monitored by optical density at 600 nm (OD). The bacterial cell culture supernatant was filtered (0.2 µm filter) and concentrated in a 50 kDa filter to 1% of the initial volume. Sialidase activity of 50 µl concentrated bacterial cell culture supernatant was measured in a total volume of 100 µl in 0,5x PBS with 0,05 µM 2-O-(p-nitrophenyl)]-α-D-N-acetylneuraminic acid and the absorbance was monitored at 405 nm after 10 minutes.

### Enterococcus faecalis, Streptococcus mitis A and Strepotoccus mitis B sialidase cloning, expression and purification

The coding sequence of Enterococcus faecalis FR191 sialidase was synthesized (Integrated DNA technologies, Belgium). The sequence harboring a C-terminal 6x-His Tag was inserted into a pET9d vector^38^ and amplified in E. coli XL1-blue. Sequencing of the plasmid was performed to verify proper insertion and the plasmid was transferred to E. coli BL21(De3) for expression. The coding sequences of *Streptococcus mitis* sialidase A and B were also harbouring a C-terminal 6x-HIS Tag and ordered already inserted in a pET9a vector (BioCat, Germany). The plasmid was amplified in E. coli XL1-blue and transferred to E. coli BL21(De3) for expression as well.

Expression for all three sialidases was induced with 1 mM IPTG for 24 h, cells were pelleted and washed with PBS. The pellets were resuspended in 20 mM TrisHCl pH 7.5, 1M NaCl supplemented with protease inhibitors (Roche), 1% Triton-X-100 and 100 mg/l lysozyme (Serva). Cells were lysed by sonification (Branson Sonifier 250) for 10×15 s on ice. Non-lysed cells were removed and the supernatant was filtered twice (0.8 µm and 0.22 µm filter) and applied to a 1ml HisTrap HP column (Cytiva) equilibrated with 20 mM TrisHCl pH 7.5 and 1M NaCl. Protein was eluted with a step gradient of 60% elution buffer (20 mM TrisHCl pH 7.5, 1 M NaCl, 0.5 M imidazole). Fractions containing the eluted proteins were pooled and desalted. Purity of the obtained proteins was verified in a polyacrylamide gel stained with Commassie and immunoblotting using the His-Tag of the recombinant protein. Finally, the purified *Enterococcus faecalis* sialidase showed a sialidase activity of 1.75 U/mg protein. Enzymatic activity of purified sialidase was measured as mentioned above using the 2-O-(p-nitrophenyl)]-α-D-N-acetylneuraminic acid assay. The sialidase activity of the purified Streptococcus sialidases were confirmed with 4-MU-Sia as the substrate and were a little higher than the activity of the *Enterococcus faecalis* sialidase.

### Lectin analysis

For immunofluorescence staining the cholangiocellular organoids cultured in MatrigelTM domes were incubated with TrypleETM Express for 5 min at 37 °C. After centrifugation at 400 rpm for 5 min cholangiocytes were seeded on glass coverslips in expansion media (Supplementary table 4). MMNK-1 cells were seeded after trypsinization on glass coverslips in DMEM containing 10 % FCS. Once cells were ∼ 70 % confluent, they were treated with E.f. sialidase (dilution in starving medium 1:100) for 2.5 h at 37 °C and 5 % CO_2_ and then fixed using 4 % PFA/PBS. Cells were incubated with the biotinylated lectins Maackia amurensis lectin (MAL-II, 0.005 mg/mL f.c., Vector Laboratories; detecting α2,3-linked sialic acid), Sambucus nigra agglutinin (SNA, 0.01 mg/mL f.c., Vector Laboratories; detecting α2,6-linked sialic acid) or Peanut agglutinin (PNA, 0.025 mg/ml f.c., Vector Laboratories; detecting β1,3-linked galactose) for one hour at room temperature in Tris buffered saline pH 7.5 with 1 mM MgCl2, 1 mM MnCl_2_ and 1 mM CaCl_2_. After washing with PBS, cells were incubated with Streptavidin-Cy3 (0.005mg/ml f.c., Rockland) and DAPI (Stock 0.6 µM, Merck) for 1 h at room temperature. Processed coverslips were mounted in Vectashield HardSetTM mounting medium (Vectorlabs). Images were taken with Leica DMi8 inverted microscope and further processed with ImageJ.

For lectin staining before and after treatment with *Streptococcus mitis* sialidase A and B, MMNK-1 cells were treated with the sialidases (diluted in starving medium 1:29) for 2.5 h at 37 °C and 5 % CO_2,_ trypsinated and washed with PBS. Lectin staining was performed with the biotinylated lectin MAL-II (0.001 mg/mL f.c., Vector Laboratories) or PNA conjugated with AlexaFluor 647 (0.001 mg/ml f.c., Vector Laboratories) for 30 minutes on ice. For MAL-II staining, cells were incubated with Streptavidin AF647 (0.0036mg/mL f.c., Jackson) for 20 minutes on ice afterwards. Cells were washed, resuspended and analyzed in a cytometer (Partec), FlowJo v10 was used for further analysis.

### Cell viability (MTT) assay

Cholangiocellular organoids cultured in MatrigelTM domes were incubated with TrypleETM Express for 5 min at 37 °C. After centrifugation at 400 rpm for 5 min cholangiocytes were seeded in a 48-well plate coated with a 7 % MatrigelTM solution in expansion medium (Extended Data Table 5/6new). After 72 h cells were treated with E.f. sialidase (0.0175 U/mg f.c.) in FCS free medium for 2.5 h at 37 °C and 5 % CO_2_. Then, cells were either incubated in experiment medium only, or in experiment media containing 750 µM GCDC for 18 hours. Then, cells were incubated with MTT solution (MTT Cell Growth Assay Kit, Merck) diluted 1:4 in starving media for 1 h at 37 °C. After washing with PBS, 100 µL isopropanol was added to each well and 80 µL of the isopropanol solution containing the blue pigment was transferred to a 96-well. Absorbance at 562 nm and 650 nm (reference) was measured using a plate reader (BioTek).

### AnnexinV Apoptosis Assay

Cholangiocellular organoids were seeded and treated as described above. After treatment with GCDC, cells were detached with TrypleE ExpressTM and stained with AnnexinV/Propidium iodide (BD Biosciences) as described in the manufacture’s instruction. Flow cytometry analysis was performed in a Cytometer (Partec) and data were analyzed with the FlowJo v10 software.

### Lectin staining of human liver biopsies

Liver biopsies from patients with PSC included in our cohort were used for lectin staining. Sections were deparaffinized and rehydrated. Antigen retrieval (Target retrieval solution pH 6, Dako Agilent) was performed before blocking of endogenous peroxidase with Peroxidase-blocking solution (Dako Agilent) and subsequent incubation with lectin blocking buffer (Roche) to prevent unspecific binding. Afterwards, sections were incubated with or without 0.175 U/mg f.c. E.f. sialidase for 2 h at 37 °C in PBS prior to incubation with lectins to prove specificity of the lectin binding. Biontinylated SNA (0.27 µg/ml f.c.) was added overnight at 4 °C diluted in Tris buffered saline pH 7.5 with 1 mM MgCl_2_, 1 mM MnCl_2_, and 1 mM CaCl_2_ to detect glycan epitopes. This was followed by incubation with streptavidin-AP (4 µg/ml f.c., Invitrogen) for 1 h at room temperature. Endogenous alkaline phosphatase was blocked using 5 mM levamisole. Vector Red (Vector Laboratories, SK5100) was used for antigen detection. Sections were then counterstained with haematoxylin and mounted in Vitro-Clud (Langenbrinck GmbH).

Images were taken with Zeiss Observer.Z1 microscope with an AxioCam MRm digital camera and further analyzed with Zeiss Zen software. Twelve different human liver biopsies were stained in which a total of 108 bile ducts were found. Bile ducts were visually inspected with focus on the apical side of the cholangiocytes.

### mRNA qPCR of sialidase genes

The mixture of 250 ul bile and 750 ul TRI-Reagent (ambion) incubated for 5 minutes at RT. 150ul Chloroform were added followed by incubation for 2-3 minutes at RT and centrifugation for 15 minutes at 12.000 x g and 4°C. Phase separation was created. The aqueous phase was transferred into a new 1,5ml tube and combined with 375ul of isopropanol. Incubation on ice for 10 minutes. Centrifugation for 10 minutes at 12.000 x g at 4°C. The supernatant was discarded. Resuspension of the pallet with 750ul of 75% ethanol. Centrifugation at 7500 x g at 4°c for 5 minutes. The supernatant was discarded and the pellet was air dried for 5-10 minutes. Resuspension with 20ul of RNase-free water. Successful extraction was confirmed by measuring the amount of RNA (ng/ul) and the 260/280nm-ratio of every sample using a NanoDrop spectrophotometer (Thermo Fisher). The resulting RNA was either stored at-80°C or transformed into cDNA immediately. Reverse transcription was conducted using the SuperScript III Reverse Transcriptase Kit (Thermo Fisher) according to the manufacturer’s instructions.

Primer sequences are listed in Extended Data Table 7. The preparation of the reaction mixture (20ul containing 10ul of the the SYBR Green Supermix (Bio-Rad), 1ul of a Primer Mix (containing 3 pairs of forward and reverse primers combined in a 1:10 ratio), 1ul of template cDNA and 8ul of nuclease-free water) was implemented on ice and protected from direct light exposure. The samples were arranged in triplicates on 96-well-plates and sealed with sealing tape for protection. qPCR cycle parameter: 2-minute period at 95°C, 40 cycles of denaturation at 95°C for 10 seconds, 1 minute annealing period at 60°C. A melting curve was recorded after every run by eventually increasing the temperature to 95°C. The resulting data was analyzed using the qPCR soft 4.1 software (analytic jena).

### Statistics

For statistical analysis and for graphic design primer 7 (PRIMER-E), Graph Pad Prism 7 (Graph Pad Software) and SPSS version 23.0.0.0 (IBM) were used. Comparison of multiple groups was performed using Kruskal-Wallis test with correction for multiple comparison using Dunn’s test or in case of viability assays using one-way ANOVA, and post-hoc Dunnett’s multiple comparison tests. Comparison of two groups was performed using either Wilcoxon matched-pairs signed rank test for paired analysis or Mann-Whitney-U test with correction for multiple comparison using Benjamini-Hochberg. Outcome analysis: one sample per patients was included. Each calculation was performed twice (with the first or the last sample). Patients with a CCA at time point of sampling were excluded (n = 8). For pairwise comparison Log Rank test (Mantel-Cox test) was used. Bilotype Definition: Relative abundance of the most abundant genus >33%.

### Ethics

The study was performed according to the World Medical Association Declaration of Helsinki and had been approved by the ethics committee of the Hannover Medical School (approval no. 220-2007, approval no. 3241-2016, approval no. 9660_BO_K_2021 and approval no. 10183_BO_K_2022).

## Data Availability

Raw data from 16S amplicon sequencing, metagenomic sequence data, bacterial genome data and data from mass spectrometry will be available upon publication after peer-review.
All additional data in the present study are available upon reasonable request to the authors.

## Extended Data

**Extended Data Table 1:** Quantitatively most important genera of the biliary microbial community in PSC patients.

| genus | prevalence (%) | mean RA±SD (%) |
| --- | --- | --- |
| Enterococcus | 100 | 23.3±33.1 |
| Streptococcus | 100 | 16.1±22.1 |
| Prevotella | 99.0 | 4.8±8.7 |
| Veillonella | 98.1 | 2.4±4.6 |
| Fusobacterium | 96.7 | 3.4±8.2 |
| Escherichia/Shigella | 96.7 | 8.7±20.6 |
| Schaalia | 94.7 | 1.4±3.3 |
| Haemophilus | 90.9 | 2.7±10.7 |
| Granulicatella | 88.5 | 1.2±3.0 |
| Bifidobacterium | 88.0 | 1.3±4.3 |
| Staphylococcus | 85.6 | 5.0±15.9 |
| Enterobacter/Pantoea | 84.2 | 4.6±14.4 |
| Klebsiella | 81.3 | 5.6±18.1 |
| Bacteroides | 80.4 | 1.5±6.1 |
| Clostridium sensu stricto | 69.4 | 1.7±7.1 |
| Pseudomonas | 29.7 | 1.0±8.1 |

n=209. grey highlighted: prevalence <80%. RA = relative abundance. SD = standard deviation

**Extended Data Table 2.**
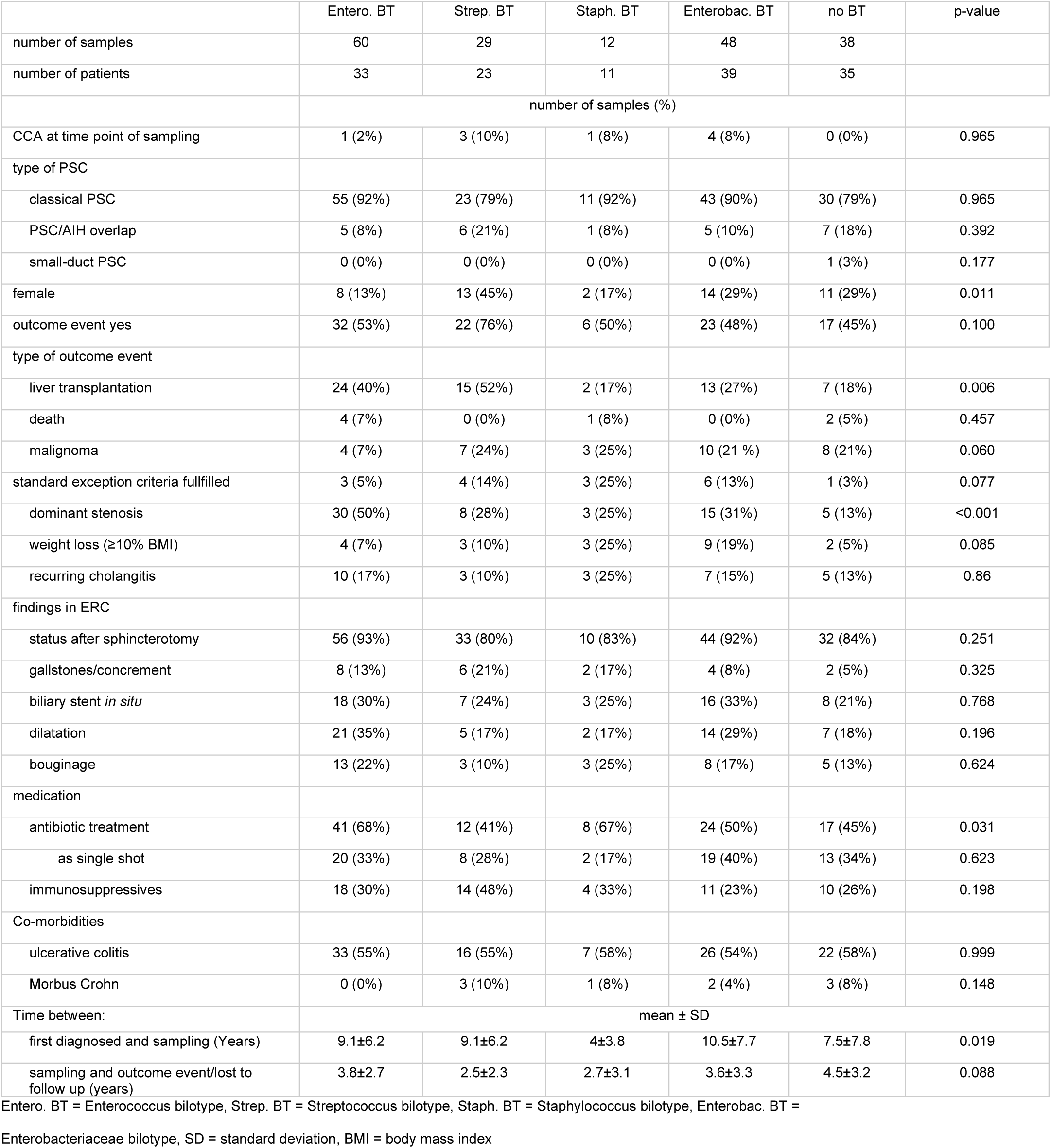
Clinical characteristics of Bilotype groups.

|  | Entero. BT | Strep. BT | Staph. BT | Enterobac. BT | no BT | p-value |
| --- | --- | --- | --- | --- | --- | --- |
| number of samples | 60 | 29 | 12 | 48 | 38 |  |
| number of patients | 33 | 23 | 11 | 39 | 35 |  |
|  | number of samples (%) |  |  |  |  |  |
| CCA at time point of sampling | 1 (2%) | 3 (10%) | 1 (8%) | 4 (8%) | 0 (0%) | 0.965 |
| type of PSC |  |  |  |  |  |  |
| classical PSC | 55 (92%) | 23 (79%) | 11 (92%) | 43 (90%) | 30 (79%) | 0.965 |
| PSC/AIH overlap | 5 (8%) | 6 (21%) | 1 (8%) | 5 (10%) | 7 (18%) | 0.392 |
| small-duct PSC | 0 (0%) | 0 (0%) | 0 (0%) | 0 (0%) | 1 (3%) | 0.177 |
| female | 8 (13%) | 13 (45%) | 2 (17%) | 14 (29%) | 11 (29%) | 0.011 |
| outcome event yes | 32 (53%) | 22 (76%) | 6 (50%) | 23 (48%) | 17 (45%) | 0.100 |
| type of outcome event |  |  |  |  |  |  |
| liver transplantation | 24 (40%) | 15 (52%) | 2 (17%) | 13 (27%) | 7 (18%) | 0.006 |
| death | 4 (7%) | 0 (0%) | 1 (8%) | 0 (0%) | 2 (5%) | 0.457 |
| malignoma | 4 (7%) | 7 (24%) | 3 (25%) | 10 (21 %) | 8 (21%) | 0.060 |
| standard exception criteria fulfilled | 3 (5%) | 4 (14%) | 3 (25%) | 6 (13%) | 1 (3%) | 0.077 |
| dominant stenosis | 30 (50%) | 8 (28%) | 3 (25%) | 15 (31%) | 5 (13%) | <0.001 |
| weight loss (≥10% BMI) | 4 (7%) | 3 (10%) | 3 (25%) | 9 (19%) | 2 (5%) | 0.085 |
| recurring cholangitis | 10 (17%) | 3 (10%) | 3 (25%) | 7 (15%) | 5 (13%) | 0.86 |
| findings in ERC |  |  |  |  |  |  |
| status after sphincterotomy | 56 (93%) | 33 (80%) | 10 (83%) | 44 (92%) | 32 (84%) | 0.251 |
| gallstones/concrement | 8 (13%) | 6 (21%) | 2 (17%) | 4 (8%) | 2 (5%) | 0.325 |
| biliary stent *in situ* | 18 (30%) | 7 (24%) | 3 (25%) | 16 (33%) | 8 (21%) | 0.768 |
| dilatation | 21 (35%) | 5 (17%) | 2 (17%) | 14 (29%) | 7 (18%) | 0.196 |
| bouginage | 13 (22%) | 3 (10%) | 3 (25%) | 8 (17%) | 5 (13%) | 0.624 |
| medication |  |  |  |  |  |  |
| antibiotic treatment | 41 (68%) | 12 (41%) | 8 (67%) | 24 (50%) | 17 (45%) | 0.031 |
| as single shot | 20 (33%) | 8 (28%) | 2 (17%) | 19 (40%) | 13 (34%) | 0.623 |
| immunosuppressives | 18 (30%) | 14 (48%) | 4 (33%) | 11 (23%) | 10 (26%) | 0.198 |
| Co-morbidities |  |  |  |  |  |  |
| ulcerative colitis | 33 (55%) | 16 (55%) | 7 (58%) | 26 (54%) | 22 (58%) | 0.999 |
| Morbus Crohn | 0 (0%) | 3 (10%) | 1 (8%) | 2 (4%) | 3 (8%) | 0.148 |
| Time between: | mean ± SD |  |  |  |  |  |
| first diagnosed and sampling (Years) | 9.1±6.2 | 9.1±6.2 | 4±3.8 | 10.5±7.7 | 7.5±7.8 | 0.019 |
| sampling and outcome event/lost to follow up (years) | 3.8±2.7 | 2.5±2.3 | 2.7±3.1 | 3.6±3.3 | 4.5±3.2 | 0.088 |

Entero. BT = Enterococcus bilotype, Strep. BT = Streptococcus bilotype, Staph. BT = Staphylococcus bilotype, Enterobac. BT =
Enterobacteriaceae bilotype, SD = standard deviation, BMI = body mass index

**Extended Data Table 3.** Top five functional pathways that differentiate best between samples with or without a disadvantageous bilotype in patient history.

| Pathway | higher mean rank in | relative abundance (%) mean±SD |  | adj. p-value | pathway description |
| --- | --- | --- | --- | --- | --- |
|  |  | disadv. BT yes | disadv. BT no |  |  |
| RHAMCAT-PWY | disadv. BT no | 0.11±0.15 | 0.35±0.25 | 6.17x10<sup>-14</sup> | L-rhamnose degradation I |
| PWY-5265 | disadv. BT yes | 0.16±0.20 | 0.04±0.09 | 1.05x10<sup>-07</sup> | peptidoglycan biosynthesis II |
| LACTOSECAT-PWY | disadv. BT yes | 0.52±0.39 | 0.19±0.26 | 4.78x10<sup>-11</sup> | lactose and galactose degradation I |
| HEMESYN2-PWY | disadv. BT yes | 0.50±0.26 | 0.30±0.17 | 3.04x10<sup>-08</sup> | heme b biosynthesis II (oxygen-independent) |
| GLCMANNANAUT-PWY | disadv. BT yes | 0.46±0.29 | 0.23±0.17 | 3.73x10<sup>-08</sup> | superpathway of N-acetylglucosamine, N-acetylmannosamine and N-acetylneuraminate degradation |

disadv. BT = disadvantageous bilotype in patient history, SD = standard deviation, adj. = adjusted, disadv. BT no: n = 142, disadv. BT yes: n = 67

**Extended Data Table 4.** seventeen bacterial pathways (MetaCyc) involved in glycocalyx components.

| pathway abbreviation | pathway |
| --- | --- |
| FUC-RHAMCAT-PWY | superpathway of fucose and rhamnose degradation |
| FUCCAT-PWY | fucose degradation |
| GALACT-GLUCUROCAT-PWY | superpathway of hexuronide and hexuronate degradation |
| GALACTARDEG-PWY | D-galactarate degradation I |
| GLCMANNANAUT-PWY | superpathway of N-acetylglucosamine |
| GLUCARDEG-PWY | D-glucarate degradation I |
| GLUCARGALACTSUPER-PWY | superpathway of D-glucarate and D-galactarate degradation |
| GLUCOSE1PMETAB-PWY | glucose and glucose-1-phosphate degradation |
| GLUCUROCAT-PWY | superpathway of β-D-glucuronosides degradation |
| LACTOSECAT-PWY | lactose and galactose degradation I |
| LIPASYN-PWY | phospholipases |
| P441-PWY | superpathway of N-acetylneuraminate degradation |
| PWY-6317 | D-galactose degradation I (Leloir pathway) |
| PWY-6713 | L-rhamnose degradation II |
| PWY-6901 | superpathway of glucose and xylose degradation |
| PWY-7242 | D-fructuronate degradation |
| RHAMCAT-PWY | L-rhamnose degradation I |

**Extended Data Table 5.** Sialidases encoded in isolates retrieved from bile samples and their comparison with salidases of validated function.

| taxonomy | accession number<br>sialidase | P62576 NanA *S.*<br>*pneumoniae* | WP_003075594 NanA<br>*S. intermedius* | WP_000739360 NanC<br>*S. pneumoniae* | Q54727 NanB *S.*<br>*pneumoniae* | WP_112091347 *F.*<br>*prausnitzii* |
| --- | --- | --- | --- | --- | --- | --- |
| *Streptococcus* sp. FR074<br>symci/mitis group | RRP56_08885 |  |  | 51 | 98 |  |
| *Streptococcus* sp. FR100<br>infantis/timonensis group | 100_09125 | 64 | 66 |  |  |  |
|  | RRP83_09055 | 69 | 63 |  |  |  |
| *Streptococcus* sp. FR109<br>infantis/timonensis group | RRP55_03930 | 64 | 66 |  |  |  |
|  | RRP55_03925 | 69 | 63 |  |  |  |
| *Streptococcus* sp. FR110<br>symci/mitis group | RRP54_07775 | 71 | 67 |  |  |  |
| *Streptococcus* sp. FR114<br>oralis group | RRP87_07470 | 71 | 66 |  |  |  |
| *Streptococcus* sp. FR158<br>infantis/timonensis group | RRQ13_05665 | 69 | 66 |  |  |  |
| *Streptococcus* sp. FR159B<br>oralis group | RRP86_07190 | 66 | 60 |  |  |  |
| *E. malodoratus* FR019 | RRA50_12200 |  |  |  |  | 56 |
| *E. gallinarum* FR047 | RRA22_07795 |  |  |  |  | 57 |
| *E. faecalis* FR191 | RG666_12025 |  |  | 76 | 48 |  |

Values are given as similarity (%). All gene sequences are available under the given accession number.

**Extended Data Table 6.** Composition of expansion medium.

| Components | Concentration |
| --- | --- |
| Advanced DMEM/F12 |  |
| GlutaMAX | 1x |
| PenStrep | 1x |
| Primocin | 100 µg/mL |
| HEPES | 10 mM |
| N2 | 1x |
| B27 | 2% |
| hEGF | 50 ng/mL |
| N-Acetyl-cysteine | 1,25 mM |
| Gastrin | 10 nM |
| Nicotinamide | 10 mM |
| Rspo1 | 1 µg/mL |
| A83-01 | 5 µM |
| Forskolin | 10 µM |
| rhFGF-10 | 100 ng/mL |
| hHGF | 25 ng/mL |
| Y-2763 | 10 µM |

**Extended Data table 7.**
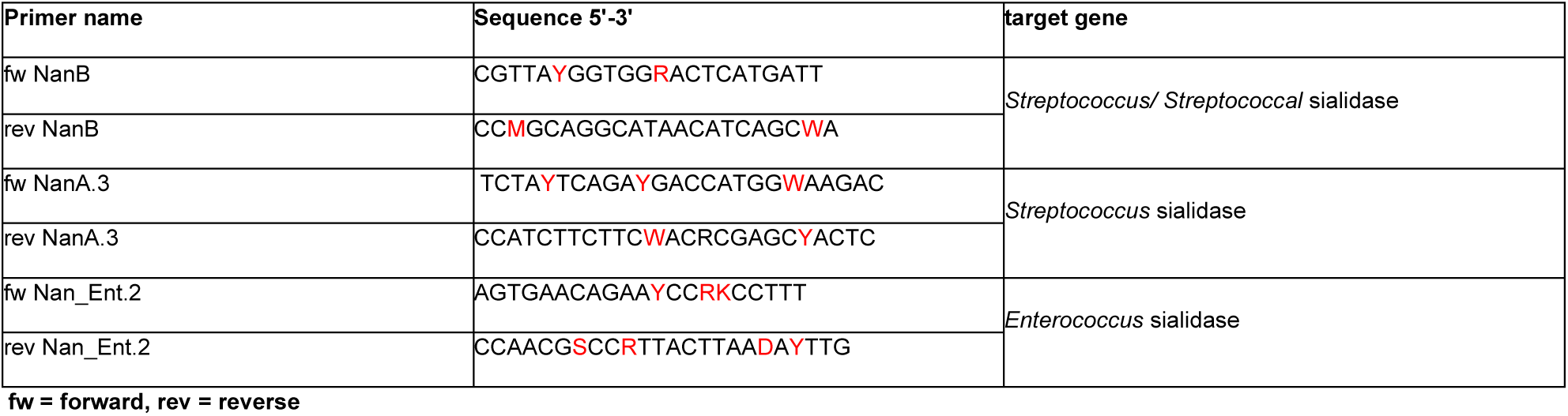
Forward and reverse primer for qPCR of sialidase genes (cDNA)

### Extended Data Figure 1

16S profile of all bile samples of the validation cohort. Each bar indicates one sample. Non-bilotype defining genera with a mean relative abundance of less than 1% are subsumed as others. n=56

### Extended Data Figure 2

SNA lectin staining of representative liver biopsy samples (immunohistochemistry, scale bar = 20 µm). Selected areas of three different biopsies/patients (p1-3) are shown exemplarily. Intensity of SNA staining and sialylation of cholangiocytes differs even within samples. Arrows: intense SNA staining in bile ducts, arrowheads: absence of SNA staining in bile ducts. Examination of 108 bile ducts in 12 liver biopsies.

### Extended Data Figure 3

Phylogenetic tree showing the relatedness of sialidases. The evolutionary history was inferred using the

Neighbor-Joining method in MEGA 7 The evolutionary distances were computed using the p-distance method and are in the units of the number of base differences per site. All ambiguous positions were removed for each sequence pair (pairwise deletion option). The numbers above branches are bootstrap support values > 50 % from 100 replications. Proteins of validated function are marked by a turquoise dot, proteins observed in the metagenomic dataset with a black dot and proteins observed in isolates from bile with a red dot.

### Extended Data Figure 4

Tree inferred with FastME 2.1.6.1 (Lefort V, Desper R, Gascuel O. FastME 2.0: A comprehensive, accurate, and fast distance-based phylogeny inference program. Mol Biol Evol. 2015;32: 2798–2800) from GBDP distances calculated from genome sequences. The branch lengths are scaled in terms of GBDP distance formula d5. The numbers above branches are GBDP pseudo-bootstrap support values > 60 % from 100 replications, with an average branch support of 72.0 %. The tree was rooted at the midpoint.

## Acknowledgements

The study was funded by the Deutsche Forschungsgemeinschaft (DFG, German Research Foundation) under Germany’s Excellence Strategy – EXC 2155 “RESIST” – Project ID 390874280 and by the German Centre for Infection Research (DZIF e.V.).

This work was supported by a grant from the German Federal Ministry of Education and Research (reference number: 01EO1302) and by the Helmholtz Association’s Initiative on Aging and Metabolic Programming. Thanks to Kirti Shukla for support in cell culture experiments, Till Robin Lesker for support in genome assemblies and Yonatan Ayalew Mekonnen for his involvement in data analysis.

## Author contributions

Friederike Klein:conception, study design, sample preparation for sequencing, bioinformatic data processing, in-silico metagenomics, statistical analysis, collection of clinical data, writing of the manuscript,

Freya Wellhöner: study design, cell culture experiments, immunofluorescence staining, sialidase activity measurements, analysis of human liver biopsies, statistical analysis, writing of the manuscript

Anika Freise: study design, organoid culture establishment, cell culture experiments, immunofluorescence staining, sialidase activity measurements, analysis of human liver biopsies, writing of the manuscript

Kristina M. Niculovic: cell culture experiments, immunofluorescence staining, sialidase activity measurements, analysis of human liver biopsies, critical reading and revising of the manuscript

Howard Junca: *in silico* metagenomics, data processing, bioinformatical analysis of metagenomics and whole genome sequences, phylogenetic analysis, critical reading and revising of the manuscript

Manuel Vicente: cell culture experiments, immunofluorescence staining, analysis of human liver biopsies, critical reading and revising of the manuscript

Elina Kats: cell culture experiments, immunofluorescence staining, sialidase activity measurements, critical reading and revising of the manuscript

Luiz Gustavo dos Anjos Borges: bioinformatical processing and analysis of metagenomic data and whole genome sequences, phylogenetic analysis, critical reading and revising of the manuscript

Leonard Knegendorf: isolation of bacteria, sialidase activity measurements, critical reading and revising of the manuscript

Karsten Cirksena: mass-spectrometry, critical reading and revising of the manuscript

Antonia Marie Triefenbach: qPCR measurements

Franziska Woelfl: organoid culture experiments, statistical analysis, critical reading and revising of the manuscript

Helin Fatma Abdullah: bioinformatical data processing, collection of clinical data, critical reading and revising of the manuscript

Meike Schulz: cell culture experiments, immunofluorescence stainings, critical reading and revising of the manuscript

Iris Plumeier: sample preparation for sequencing, isolation of bacteria, critical reading and revising of the manuscript

Silke Kahl: sample preparation for sequencing, bioinformatical data processing, critical reading and revising of the manuscript

Iris Albers: staining of human liver biopsies, critical reading and revising of the manuscript

Martijn Zoodsma: machine learning analysis, critical reading and revising of the manuscript

Marius Vital: bioinformatical data processing, critical reading and revising of the manuscript

Torsten Voigtländer: bile sampling, clinical advisor, critical reading and revising of the manuscript

Henrike Lenzen: bile sampling, clinical advisor, critical reading and revising of the manuscript

Jessica Schmitz: staining, digitization and morphological evaluation of human liver biopsies, critical reading and revising of the manuscript

Anna Saborowski: cell culture experiments, critical reading and revising of the manuscript

Michael P. Manns: conception, study design, critical reading and revising of the manuscript

Philipp Solbach: conception, study design, critical reading and revising of the manuscript

Jan Hinrich Bräsen: tissue sample selection, analysis of human liver biopsies, critical reading and revising of the manuscript

Gisa Gerold: mass-spectrometry, critical reading and revising of the manuscript

Cheng-Jian Xu: machine learning analysis, critical reading and revising of the manuscript

Heiner Wedemeyer: conception, writing of the manuscript, critical reading and revising of the manuscript

Anja K. Münster-Kühnel: study design, cell culture experiments, immunofluorescence staining, sialidase activity measurements, analysis of human liver biopsies, writing of the manuscript

Dietmar H. Pieper: conception, study design, sequencing, bioinformatical data analysis, phylogenetic analysis, isolation of bacteria, writing of the manuscript

Benjamin Heidrich: conception, study design, in silico metagenomics, bioinformatical data analysis, statistical analysis, writing of the manuscript

## Data Availability

Raw data from 16S amplicon sequencing, metagenomic sequence data, bacterial genome data and data from mass spectrometry will be available upon publication after peer-review.

## Abbreviations

adj-p: adjusted p-value
AUC: Area under receiver-operator characteristics
BMI: body mass index
BT: bilotype
CCA: cholangiocellular adenocarcinoma
disadv.: disadvantageous
E.f.: *Enterococcus faecalis* sialidase
Enterobact.: Enterobacteriaceae
ERC: endoscopic retrograde cholangiography
fw: Forward
GCDC: glycochenodeoxycholic acid
MAL: *Maackia amurensis* lectin
MRI: magnetic resonance imaging
PNA: Peanut agglutinin
PSC: primary sclerosing cholangitis
qPCR: quantitative PCR
RA: relative abundance
rev: reverse
RKPM: reads per kilo base per million mapped reads
SD: standard deviation
SNA: *Sambucus nigra* agglutinin

## Main references

1. Isayama H, Tazuma S, Kokudo N, Tanaka A, Tsuyuguchi T, Nakazawa T, et al. Clinical guidelines for primary sclerosing cholangitis 2017. J Gastroenterol. 2018;53(9):1006–1034.

2. Karlsen TH, Folseraas T, Thorburn D, Vesterhus M. Primary sclerosing cholangitis-a comprehensive review. J Hepatol. 2017;67(6):1298–1323.

3. Maillette de Buy Wenniger, L J, Hohenester S, Maroni L, van Vliet SJ, Oude Elferink RP, Beuers U. The cholangiocyte glycocalyx stabilizes the ‘biliary HCO3 umbrella’: An integrated line of defense against toxic bile acids. Dig Dis. 2015;33(3):397–407.

4. Hohenester S, Wenniger LM, Paulusma CC, van Vliet SJ, Jefferson DM, Elferink RP, et al. A biliary HCO3-umbrella constitutes a protective mechanism against bile acid-induced injury in human cholangiocytes. Hepatology. 2012;55(1):173–183.

5. Liwinski T, Zenouzi R, John C, Ehlken H, Ruhlemann MC, Bang C, et al. Alterations of the bile microbiome in primary sclerosing cholangitis. Gut. 2020;69(4):665–672.

6. Hirschfield GM, Karlsen TH, Lindor KD, Adams DH. Primary sclerosing cholangitis. Lancet. 2013;382(9904):1587-1599.

7. Trivedi PJ, Bowlus CL, Yimam KK, Razavi H, Estes C. Epidemiology, natural history, and outcomes of primary sclerosing cholangitis: A systematic review of population-based studies. Clin Gastroenterol Hepatol. 2022;20(8):1687–1700.e4.

8. Keitel V, Reich M, Haussinger D. TGR5: Pathogenetic role and/or therapeutic target in fibrosing cholangitis? Clin Rev Allergy Immunol. 2015;48(2-3):218–225.

9. Allegretti JR, Kassam Z. Fecal microbiota transplantation in patients with primary sclerosing cholangitis: The next steps in this promising story. Am J Gastroenterol. 2019;114(8):1354–1355.

10. Hata H, Natori T, Mizuno T, Kanazawa I, Eldesouky I, Hayashi M, et al. Phylogenetics of family enterobacteriaceae and proposal to reclassify escherichia hermannii and salmonella subterranea as atlantibacter hermannii and atlantibacter subterranea gen. nov., comb. nov. Microbiol Immunol. 2016;60(5):303–311.

11. Zimmer CL, von Seth E, Buggert M, Strauss O, Hertwig L, Nguyen S, et al. A biliary immune landscape map of primary sclerosing cholangitis reveals a dominant network of neutrophils and tissue-resident T cells. Sci Transl Med. 2021;13(599):eabb3107. doi: 10.1126/scitranslmed.abb3107.

12. Rath S, Heidrich B, Pieper DH, Vital M. Uncovering the trimethylamine-producing bacteria of the human gut microbiota. Microbiome. 2017;5(1):54–9.

13. Human Microbiome Project Consortium. Structure, function and diversity of the healthy human microbiome. Nature. 2012;486(7402):207-214.

14. Douglas GM, Maffei VJ, Zaneveld JR, Yurgel SN, Brown JR, Taylor CM, et al. PICRUSt2 for prediction of metagenome functions. Nat Biotechnol. 2020;38(6):685–688.

15. Caspi R, Altman T, Billington R, Dreher K, Foerster H, Fulcher CA, et al. The MetaCyc database of metabolic pathways and enzymes and the BioCyc collection of pathway/genome databases. Nucleic Acids Res. 2014;42(Database issue):459.

16. Ji S, Juran BD, Mucha S, Folseraas T, Jostins L, Melum E, et al. Genome-wide association study of primary sclerosing cholangitis identifies new risk loci and quantifies the genetic relationship with inflammatory bowel disease. Nat Genet. 2017;49(2):269–273.

17. Folseraas T, Melum E, Rausch P, Juran BD, Ellinghaus E, Shiryaev A, et al. Extended analysis of a genome-wide association study in primary sclerosing cholangitis detects multiple novel risk loci. J Hepatol. 2012;57(2):366–375.

18. Trauner M, Fickert P, Halilbasic E, Moustafa T. Lessons from the toxic bile concept for the pathogenesis and treatment of cholestatic liver diseases. Wien Med Wochenschr. 2008;158(19-20):542–548.

19. Tabibian JH, Weeding E, Jorgensen RA, Petz JL, Keach JC, Talwalkar JA, et al. Randomised clinical trial: Vancomycin or metronidazole in patients with primary sclerosing cholangitis-a pilot study. Aliment Pharmacol Ther. 2013;37(6):604–612.

20. Rath S, Rud T, Karch A, Pieper DH, Vital M. Pathogenic functions of host microbiota. Microbiome. 2018;6(1):174–0.

21. Grobe B, Solbach P, Wellhöner F, Plumeier I, Heidrich B, Pieper DH, et al. Next generation sequencing outperforms standard microbiological cultivation for detection of bacteria in the biliary tract in patients with ischemic type biliary lesions and/or anastomotic stricture.. 2019;70(Suppl 1):e558.

22. Venkatesh SK, Welle CL, Miller FH, Jhaveri K, Ringe KI, Eaton JE, et al. Reporting standards for primary sclerosing cholangitis using MRI and MR cholangiopancreatography: Guidelines from MR working group of the international primary sclerosing cholangitis study group. Eur Radiol. 2022;32(2):923–937.

23. Chapman MH, Thorburn D, Hirschfield GM, Webster GGJ, Rushbrook SM, Alexander G, et al. British society of gastroenterology and UK-PSC guidelines for the diagnosis and management of primary sclerosing cholangitis. Gut. 2019;68(8):1356–1378.

24. Klein F, Wellhoner F, Plumeier I, Kahl S, Chhatwal P, Vital M, et al. The biliary microbiome in ischaemic-type biliary lesions can be shaped by stenting but is resilient to antibiotic treatment. Liver Int. 2022;42(5):1070–1083.

25. Callahan BJ, McMurdie PJ, Rosen MJ, Han AW, Johnson AJA, Holmes SP. DADA2: High-resolution sample inference from illumina amplicon data. Nat Methods. 2016;13(7):581–583.

26. Wang Q, Garrity GM, Tiedje JM, Cole JR. Naive bayesian classifier for rapid assignment of rRNA sequences into the new bacterial taxonomy. Appl Environ Microbiol. 2007;73(16):5261–5267.

27. Wick RR, Judd LM, Gorrie CL, Holt KE. Completing bacterial genome assemblies with multiplex MinION sequencing. Microb Genom. 2017;3(10):e000132.

28. Gurevich A, Saveliev V, Vyahhi N, Tesler G. QUAST: Quality assessment tool for genome assemblies. Bioinformatics. 2013;29(8):1072–1075.

29. Brettin T, Davis JJ, Disz T, Edwards RA, Gerdes S, Olsen GJ, et al. RASTtk: A modular and extensible implementation of the RAST algorithm for building custom annotation pipelines and annotating batches of genomes. Sci Rep. 2015;5:8365.

30. Meier-Kolthoff JP, Goker M. TYGS is an automated high-throughput platform for state-of-the-art genome-based taxonomy. Nat Commun. 2019;10(1):2182–3.

31. Meier-Kolthoff JP, Carbasse JS, Peinado-Olarte RL, Goker M. TYGS and LPSN: A database tandem for fast and reliable genome-based classification and nomenclature of prokaryotes. Nucleic Acids Res. 2022;50(D1):D801–D807.

32. Meier-Kolthoff JP, Auch AF, Klenk H, Goker M. Genome sequence-based species delimitation with confidence intervals and improved distance functions. BMC Bioinformatics. 2013;14:60–60.

33. Li J, Jia H, Cai X, Zhong H, Feng Q, Sunagawa S, et al. An integrated catalog of reference genes in the human gut microbiome. Nat Biotechnol. 2014;32(8):834–841.

34. Kuhn M. Building predictive models in R using the caret package. Journal of Statistical Software. 2008;28(5):1–26.

35. Toyoda S, Eto Y, Aoki K. Bile lysosomal enzymes: Characteristics and pathological significance for various hepatobiliary disorders. Clin Chim Acta. 1977;79(2):291–298.

36. Tyanova S, Temu T, Cox J. The MaxQuant computational platform for mass spectrometry-based shotgun proteomics. Nat Protoc. 2016;11(12):2301–2319.

37. Karen C. Carroll, Michael A. Pfaller, Marie L. Landry, et al. Manual of clinical microbiology. 12th ed. ASM Press; 2019:11-21.

38. Christensen S, Egebjerg J. Cloning, expression and characterization of a sialidase gene from arthrobacter ureafaciens. Biotechnol Appl Biochem. 2005;41(Pt 3):225–231.

