## Supplementary figures and images for "Cholangiocyte glycocalyx degradation boosts primary sclerosing cholangitis"

### Extended Data Figure 1

Extended Data Figure 1

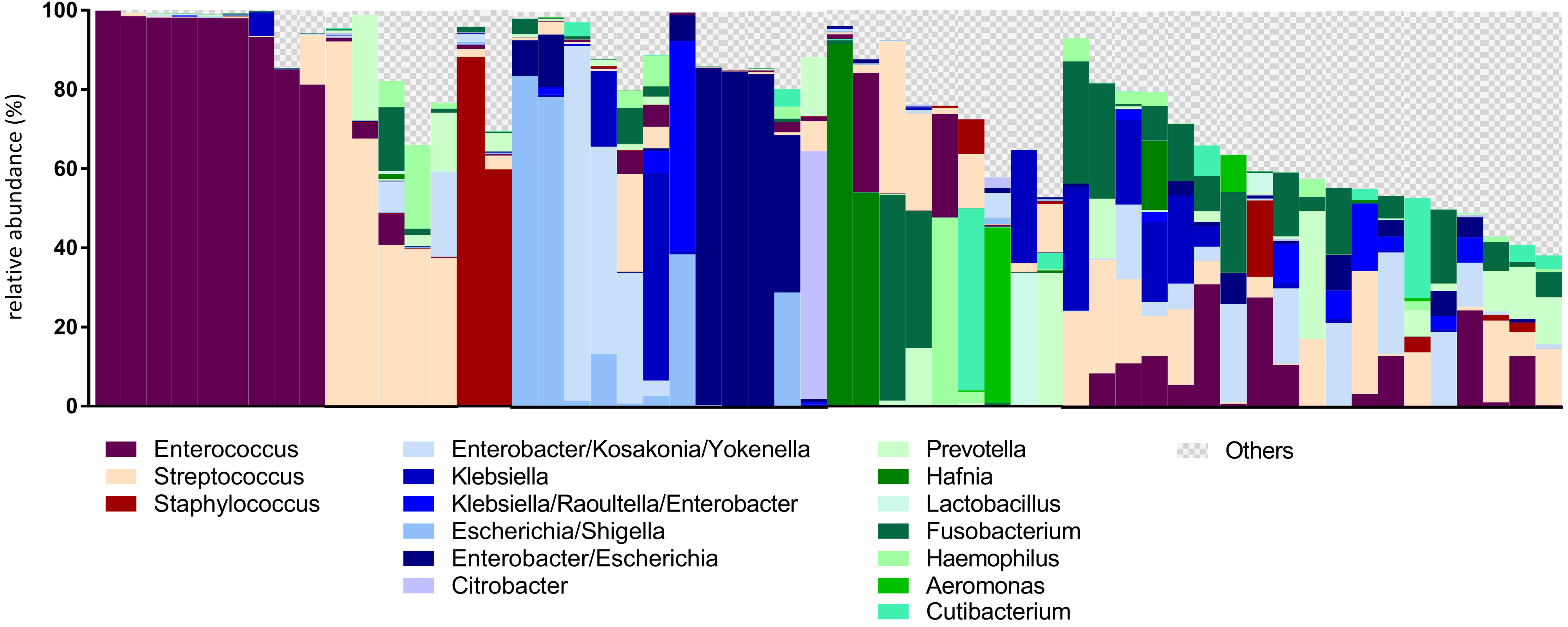

### Extended Data Figure 2

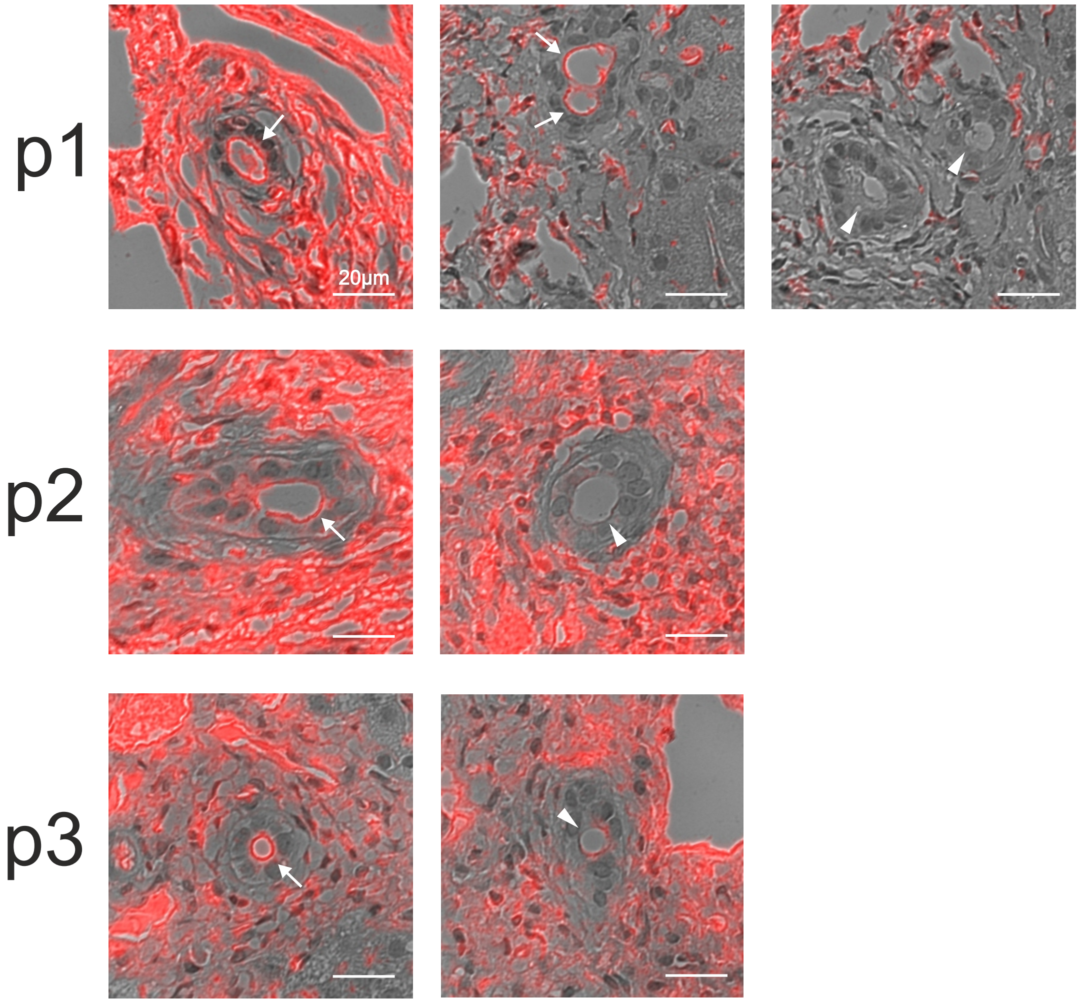


Extended Data Figure 2

### Extended Data Figure 3

Extended Data Figure 3

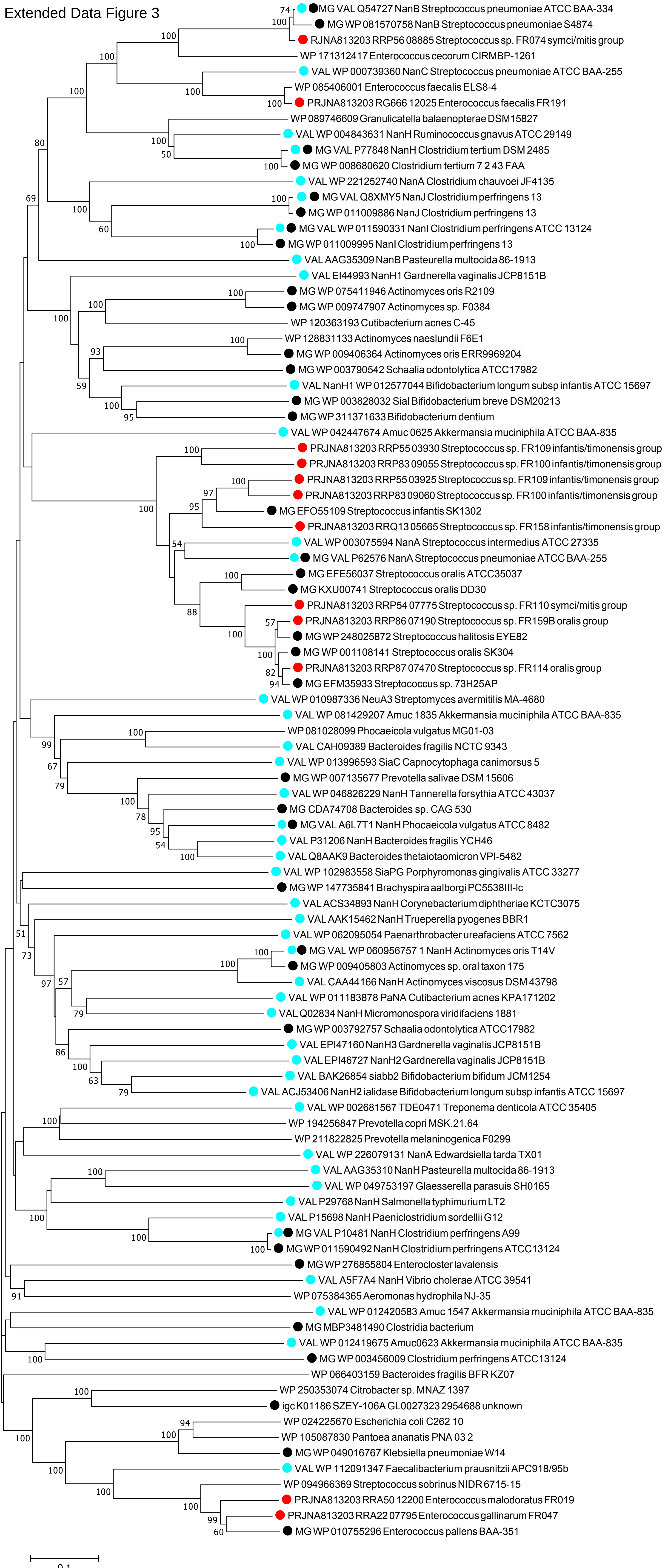

### Extended Data Figure 4

Extended Data Figure 4

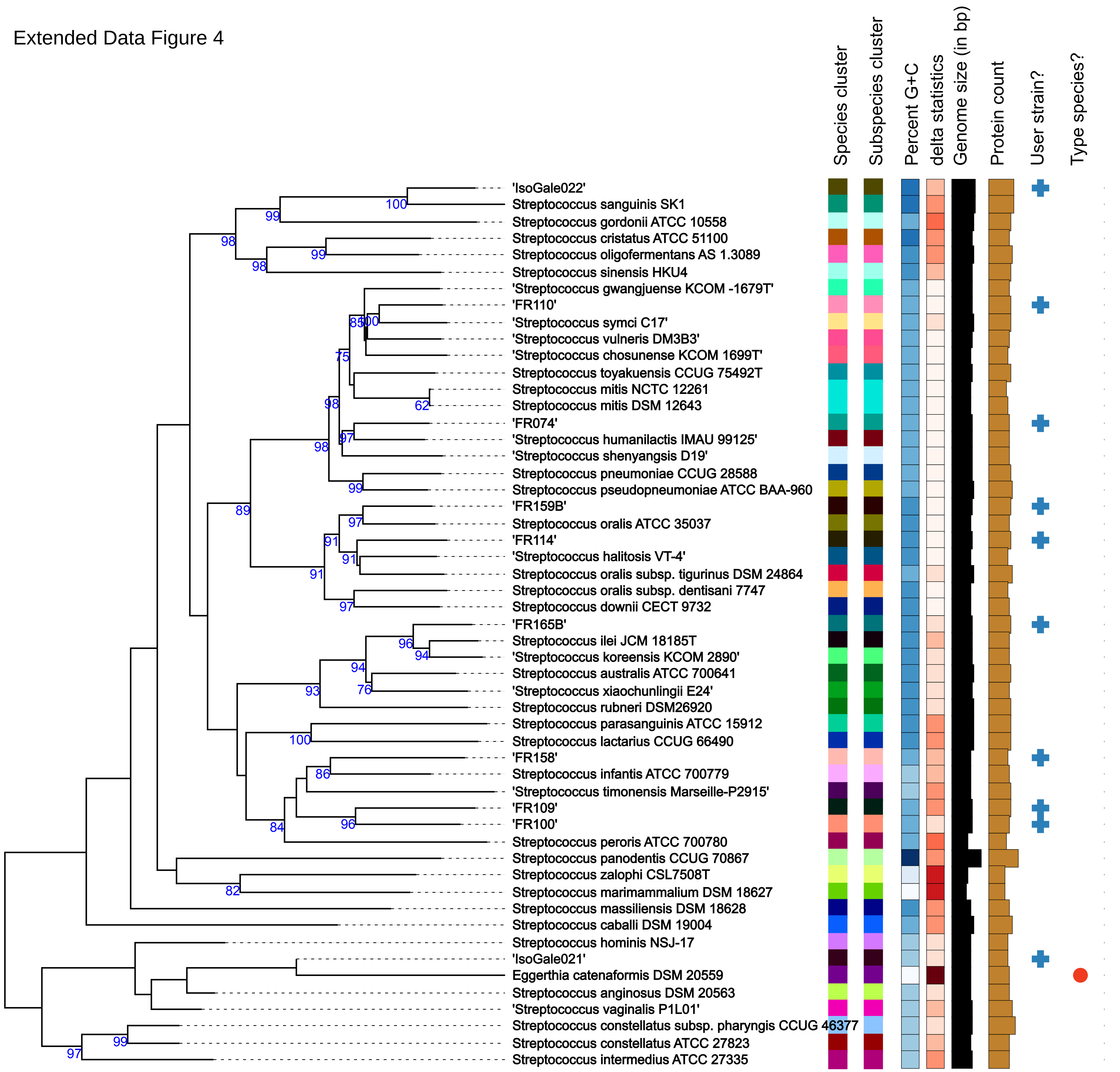
